# Testing Bidirectional Associations Between Screen Time and Inattention/Hyperactivity Symptoms From Childhood to Adulthood in a Brazilian Cohort

**DOI:** 10.1101/2025.07.26.25332224

**Authors:** Ricardo Kaciava Bombardelli, Patricia Pinheiro Bado, João Pedro Gonçalves Pacheco, Gabriele dos Santos Jobim, Ary Gadelha de Alencar Araripe Neto, Euripedes Constantino Miguel, Rodrigo Affonseca Bressan, Pedro Mario Pan, Luis Augusto Rohde, Giovanni Abrahão Salum, Mauricio Scopel Hoffmann

**Affiliations:** Mental Health Epidemiology Group (MHEG), Universidade Federal de Santa Maria, Santa Maria, Brazil; School of Medicine, Universidade Federal de Santa Maria, Santa Maria, Brazil; Graduate Program in Psychiatry and Behavioral Sciences, Universidade Federal do Rio Grande do Sul, Porto Alegre, Brazil; National Center for Innovation and Research in Mental Health (CISM), São Paulo, Brazil; Department of Psychology, Pontifical Catholic University of Rio de Janeiro (PUC-Rio), Rio de Janeiro, Brazil; Department of Neuropsychiatry, Universidade Federal de Santa Maria, Santa Maria, Brazil; Department of Psychiatry, Escola Paulista de Medicina, Universidade Federal de São Paulo (UNIFESP), São Paulo, Brazil; Universidade de São Paulo (USP), São Paulo, Brazil; Laboratório Interdisciplinar de Neurociências Clínicas, Departamento de Psiquiatria, Universidade Federal de São Paulo (UNIFESP), São Paulo, Brazil; Department of Psychiatry and Legal Medicine, Universidade Federal do Rio Grande do Sul, Porto Alegre, Brazil; Child Mind Institute, New York, USA; Care Policy and Evaluation Centre, London School of Economics and Political Science, London, UK

**Keywords:** Inattention, hyperactivity, screen time, children, RI-CLPM, LMIC

## Abstract

**Background:** Screen time has been linked with inattention/hyperactivity symptoms, but studies often do not distinguish within-from between-person associations over long developmental periods.

**Methods:** Data were drawn from the Brazilian High-Risk Cohort for Mental Health Conditions, a school-based cohort with assessments at baseline (2010) and follow-ups in 2013–2014 and 2018–2019, 76% retention. The sample included 2,511 children (mean age = 10.2 years at baseline, 54.7% males). Daily screen time (hours spent exposed to computer, television or video games) was obtained by asking the parent or the primary caregiver (94.9% mothers). Inattention/hyperactivity symptoms were assessed with the Strengths and Difficulties Questionnaire (SDQ). Analysis was carried out using random intercept cross-lagged panel models, adjusting for sample representativeness, attrition, and demographic covariates. Sensitivity analysis was carried out using the Attention Scale of the Child and Behavior Checklist (CBCL), harmonized with the Adult Behavior Checklist (ABCL).

**Results:** We found no within-person relationship between screen time and inattention/hyperactivity symptoms from childhood to early adulthood. Significant between-person associations were found in the SDQ but not in the adjusted SDQ models (respectively: β = 0.25; 95%CI = 0.02, 0.45; p = 0.029, and β = 0.22; 95%CI = -0.00, 0.45; p = 0.052). For instance, the history of primary caregiver’s psychiatric diagnoses was associated with higher average screen time (β = 0.16; 95% CI, 0.07, 0.25; p < 0.001) and inattention/hyperactivity symptoms (β = 0.27; 95% CI, 0.21, 0.34; p < 0.001). Findings were replicated in the CBCL-ABCL model.

**Conclusion:** We found no longitudinal association between increasing screen time exposure beyond an individual’s average and inattention/hyperactivity symptomatology. Furthermore, between-person associations were absent after covariate adjustment and results weren’t sensitive to the questionnaire.

## Introduction

The relationship between screen time and inattention/hyperactivity symptomatology has been explored in recent years, with the majority of studies being cross-sectional (Thorell, Burén, Wiman, Sandberg & Nutley, 2022). This approach limits conclusions about causation. A recent systematic literature review (Thorell et al., 2022) found from absence to bidirectional associations when longitudinal studies are examined. Moreover, most of those longitudinal studies applied models that only examine whether children with higher inattention/hyperactivity symptoms tend to have higher screen time on average (i.e., between-person effects) rather than tracking changes within the same person over time (i.e., within-person effects). It is also important to estimate within-person effects as this would provide information on what would happen to inattention/hyperactivity symptoms if screen time increases beyond what is expected for a given person, as, for example, in a pandemic scenario (Trott, Driscoll, Iraldo & Pardhan, 2022).

To address these limitations, random intercept cross-lagged panel models (RI-CLPM) are used to separate within-from between-person associations, providing a more detailed analysis of stable trait-like differences across individuals (between-person association) and state-like changes within individuals over time (within-person association) (see more at Orth 2021). However, recent RI-CLPM studies on screen time and inattention/hyperactivity come mainly from high-income countries (HICs), without assessing socioeconomic factors in different contexts such as low- and middle-income countries (LMICs). For instance, evidence suggests that higher screen time use is more prevalent among children from lower socioeconomic backgrounds (Takahashi et al., 2023). However, this is relative to what this background means in HICs. The lower class in LMICs has less access to assets such as electronic devices and the internet than HICs (Kharas, 2010; World Bank Group, 2024).

Therefore, as social background can confound the relationships of screen exposition and inattention/hyperactivity, this association might be different in a LMIC context.

Previous studies often focused on narrow age ranges, excluding the transition from childhood to adulthood, and findings on within-person associations remain inconsistent. Some studies found no link between general screen time and inattention/hyperactivity (Beyens, Piotrowski & Valkenburg, 2020; Deng et al., 2024), while others found early externalizing/inattention symptoms predicted later screen use (Neville et al., 2021). Certain screen modalities—like gaming, social media, and media multitasking—were sometimes linked to later inattention/hyperactivity (Baumgartner et al., 2017; Boer et al., 2020; Tiraboschi et al., 2025), though reverse effects also emerged (Beyens et al., 2020; Tiraboschi et al., 2025). Still, several within-person studies found no consistent associations, or identified sex-specific patterns (Beeres et al., 2021; Beyens et al., 2020; Boer et al., 2020; Deng et al., 2024). All used short follow-up periods (≤2 years), leaving a gap in long-term, developmental research. Moreover, few studies conducted sensitivity analyses on measurement tools, despite known variability in inattention/hyperactivity estimates across instruments (Polanczyk et al., 2015), which may partly explain prior inconsistencies.

Thus, our study aims to address these limitations by examining the relationship of overall screen time (TV, computer, and video games) and inattention/hyperactivity symptoms throughout development (childhood to young adulthood) in a school-based sample from Brazil, while disentangling within-from between-person associations. We hypothesize that screen time and inattention/hyperactivity symptoms will present a between-person association (i.e., higher screen time average will be correlated with higher inattention/hyperactivity symptoms average) and that oscillations from these tendencies will not be associated (i.e., higher screen time use than expected will not increase inattention/hyperactivity symptoms at follow-ups and vice versa). We also expect that average inattention/hyperactivity symptoms and screen time will be positively related to lower socioeconomic background and higher primary caregiver’s psychopathology.

## Methods

### Sample

We analyzed baseline (2010-2011), 3-(2013-2014) and 8-year follow-ups (2018-2019) from the Brazilian High Risk Cohort study (BHRC), a large school-based cohort oversampled for high family risk for psychopathology (Salum et al., 2015). Briefly, families were recruited from 57 state-funded schools in Porto Alegre and São Paulo on the day of enrolment in 2010, which is compulsory in Brazil. A total of 8,012 caregivers (87.3% mothers) were screened by lay interviewers using a modified Family History Screen based on DSM-IV criteria to assess psychiatric conditions, and a family liability index was calculated (details can be found at Salum et al., 2015). Two subgroups were recruited: a high-risk group based on the family liability index (n=1,553) and a random sample of eligible children (n=958). Thus, the final sample of 2,511 children and adolescents underwent full assessments by lay interviewers (parent) and trained psychologists (child) at baseline (6 to 14 years of age), 2,010 at first (80% retention, 9-17 years of age) and 1,917 at the second follow-up (76.3% retention, 14-23 years of age).

### Screen time measure

Daily screen time (e.g. time spent in front of television, computer or video games) was obtained by asking the parent or the primary caregiver "How many hours a day does the child spend in front of the television, using the computer and playing video games on average"? Responses were given in total numbers of hours and classified as 1, 2, 3, 4, 5, 6 and 7, and 8 or more hours a day.

### Inattention/hyperactivity symptoms measurement and modeling

Inattention/hyperactivity symptoms were measured using the Strengths and Difficulties Questionnaire (SDQ), a 25-item parent-reported screener for emotional and behavioral problems over the past 6 months, with Likert-type responses (0=Not true; 1=Somewhat true, 2=Certainly True) (Goodman, 1997; Fleitlich-Bilyk & Goodman, 2004). A total score for inattention/hyperactivity was calculated by summing the five corresponding items, resulting in a score range of 0 to 10. Internal consistency and reliability of this dimension in our data was estimated using a SDQ four-factor model considering the wave of data collection as a cluster, as described below. For sensitivity analysis, inattention/hyperactivity symptoms were also computed using the Attention Problems scale from the Child Behavior Checklist (CBCL) harmonized with the Adult Behavior Checklist (ABCL) (8 items, scoring 0 to 16). CBCL-ABCL is also a Likert-type scale (0=Not true; 1=Somewhat true, 2=Certainly True), assessing symptoms over the past six months (Achenbach & Rescorla, 2001) (see Supplementary Methods).

### Covariates

To account for important confounders between screen time and inattention/hyperactivity symptoms, we adjusted the analysis for sex (female as reference), age, social class, maternal education, and the primary caregiver’s history of psychiatric diagnosis at baseline. Age was measured in years and standardized (mean-centered). Social class was classified by the Brazilian Association of Research Companies (ABEP, 2009). The ABEP is a parent-reported asset-based assessment that also includes the head of the household’s highest level of education and sanitation to classify subjects as belonging to the A (highest) to E (lowest and reference category) socioeconomic class. This corresponds to an average gross monthly household income of $4.937 (A), $2.686-$1.394 (B), $834-$571 (C) to $400 (D/E), according to 2014 data and currency conversion rates (ABEP, 2009). Moreover, we included maternal education, classified according to the International Standard Classification of Education 2011 as ‘Incomplete Primary or Lower Secondary’, ‘Complete Lower Secondary’, ‘Complete Upper Secondary’ and ‘Bachelor degree or equivalent’. History of psychiatric disorder in the primary caregiver (94.9% mothers) was assessed using the Mini International Psychiatric Interview (MINI), and included modules for major depressive episode, manic episode, panic disorder, agoraphobia, social anxiety disorder, alcohol abuse and dependence, drug abuse and dependence, psychotic disorders, generalized anxiety disorder and ADHD.

### Weights

The BHRC used an oversampling procedure, as described in Salum et. al, 2015. A sampling weight was generated to account for this high-risk stratification, which fully accounted for the oversampling procedure (Martel et al., 2017). In a previous analysis of the BHRC, any baseline history of psychiatric diagnosis, maternal education, presence of a psychiatric diagnosis in the primary caregiver, and study site predicted response at follow-up (Amaral et al., 2025). These four variables were used to compute inverse probability weight (IPW) to address sample attrition in all analyses (Seaman & White, 2013), as described elsewhere (Amaral et al., 2025). Sampling weight and IPW were multiplied, and weights for 36 participants were trimmed for the 1^st^ and 99^th^ percentiles. The resulting weights were used throughout the analyses to minimize bias associated with oversampling for high-risk procedures and missing data at follow-up, so that the sample is more representative of the baseline sample.

### Statistical analysis and structural equation modeling

Statistical analyses were carried out from August 15 2024 to February 28 2025. The relationship between screen time and inattention/hyperactivity symptoms was modeled using RI-CLPM across baseline (2010), first (2013-2014) and second follow-ups (2018-2019). The RI-CLPM was specified according to Orth et al. (2021) and the analysis code with model specification can be found in supporting information (Code and Statistical Analysis Syntax). Briefly, we included two random intercepts, fixing the factor loadings for each screen time and hyperactivity/inattention into 1, to account for stable, trait-like differences between individuals. These intercepts were allowed to covary, capturing any baseline associations between attentional difficulties and screen time. The within-person component captures variation and represents changes from one’s own mean level of inattention/hyperactivity symptoms as a function of changes in one’s own level of screen time use (and vice-versa). To examine within-person dynamics, we created latent variables by centering the observed variables around each individual’s mean. Autoregressive paths were specified to capture the stability of attentional difficulties and screen time over time, while cross-lagged paths were included to investigate potential reciprocal effects between the two variables. An important advantage of modeling within-person change is that it allows each individual to be their own comparator, eliminating the need to account for stable characteristics that don’t change over time, such as sex, developmental stage or socioeconomic background, which vary between different people but not as much as within the same individual.

The RI-CLPM was implemented with full information maximum likelihood as the estimator method. The product of sampling and attrition weights was included as weights in all models. Coefficients’ significance was evaluated on the basis of p < 0.05 and 95% CIs. Model goodness of fit was evaluated using root-mean-square error of approximation (RMSEA), comparative fit index (CFI), Tucker-Lewis index (TLI), and Standardized Root Mean Square Residual (SRMR). RMSEA lower than 0.060 and CFI or TLI values higher than 0.950 indicate a good-to-excellent model. SRMR lower than or equal to 0.100 indicates adequate fit, and lower than 0.060 in combination with previous indices indicates good fit (Hu & Bentler, 1999). RI-CLPMs were estimated using the *lavaan* package, implemented in R (Rosseel, 2012) We estimated two models. First, a RI-CLPM was estimated using the screen time and SDQ hyperactivity/inattention scores only. Second, the random intercepts of screen time and inattention/hyperactivity symptoms were regressed on the covariates (sex, age, social class, maternal education and primary caregiver’s history of psychiatric diagnosis at baseline) to investigate if the average screen time use and inattention/hyperactivity symptoms are confounded by these variables.

Confirmatory factor analysis (CFA) was used to estimate the SDQ four-factor model and the CBCL-ABCL 8-factor model so internal consistency and reliability indices could be estimated (Supplementary Methods; Supplementary Results). We also tested longitudinal measurement invariance of the SDQ and CBCL-ABCL models to ensure the measurement stability over time. CFA was performed using Mplus version 8.6 (Muthén & Muthén, 2017) and implemented in RStudio version 2024.04.2+764 and R version 4.4.1 using the *MplusAutomation* package (Hallquist & Wiley, 2018), which was also used to extract factor scores generated in Mplus. All bifactor reliability indices were calculated using the *BifactorIndicesCalculator* package in R (Dueber, 2017). For procedures and results, see the supporting information (Supplementary Methods; Supplementary Results; Table S1; Table S2; Table S3).

Descriptive analyses were conducted to summarize sample demographics and characteristics such as children’s age, sex, screen time, inattention/hyperactivity scores, and other covariates by wave. The Shapiro–Wilk test was used to assess the normality of inattention/hyperactivity scores, while Wilcoxon signed-rank tests were applied to evaluate significant differences across waves. All analyses were performed in RStudio (version 2024.04.2+764) using R (version 4.4.1). Descriptive statistics were generated and grouped using functions applied to the *table1* package (Rich, 2025).

### Sensitivity analysis

We estimated an additional adjusted RI-CLPM to test if results were sensitive to the inattention/hyperactivity symptoms questionnaire by replacing the SDQ-derived hyperactivity/inattention scores with the CBCL-ABCL scores.

## Results

### Descriptive statistics and sample characteristics

Sample characteristics by wave are presented in Table 1. At baseline, the study included 2,511 children, with the majority being boys (54.8%) and belonging to ABEP stratum C (middle class) across all time points. Only 37.2% of mothers had complete upper secondary education and 29.5% of the primary caregivers had a history of psychiatric diagnosis at baseline. Children had an overall daily mean screen time of 3.7 hours, SDQ inattention/hyperactivity mean score of 4.3 and CBCL-ABCL inattention/hyperactivity mean score of 3.4. Results for Shapiro-Wilk tests for normality and Wilcoxon signed-rank tests for significance differences of inattention/hyperactivity scores across waves can be found in supporting information (Supplementary Results; Table S4).

**Table 1.** Sample Demographic and Data Characteristics by Wave.

| Characteristics | Wave |  |  |  |
| --- | --- | --- | --- | --- |
|  | Baseline | Wave 1 | Wave 2 | Overall |
|  | 2010 (N=2511) | 2013 (N=2010) | 2018 (N=1905) | (N=6426) |
| Age (in years) |  |  |  |  |
| Mean (SD) | 10 (1.9) | 13 (1.9) | 18 (2.0) | 14 (3.8) |
| Sex |  |  |  |  |
| Female | 1136 (45.2%) | 885 (44.0%) | 872 (45.8%) | 2893 (45.0%) |
| Male | 1375 (54.8%) | 1125 (56.0%) | 1033 (54.2%) | 3533 (55.0%) |
| Maternal Education Level |  |  |  |  |
| Incomplete Primary<br>or Lower Secondary | 607 (24.2%) | 638 (31.7%) | 521 (27.3%) | 1766 (27.5%) |
| Complete Lower<br>Secondary | 857 (34.1%) | 497 (24.7%) | 412 (21.6%) | 1766 (27.5%) |
| Complete Upper<br>Secondary | 934 (37.2%) | 740 (36.8%) | 694 (36.4%) | 2368 (36.9%) |
| Bachelor degree or<br>equivalent | 85 (3.4%) | 86 (4.3%) | 113 (5.9%) | 284 (4.4%) |
| PCPD |  |  |  |  |
| Don't have | 1770 (70.5%) |  |  |  |
| Have (any) | 741 (29.5%) |  |  |  |
| Socioeconomic Class |  |  |  |  |

**Table 1** continued
|  |  |  |  |  |
| --- | --- | --- | --- | --- |
| A and B | 295 (11.7%) | 200 (10.0%) | 285 (15.0%) | 780 (12.1%) |
| C | 1788 (71.2%) | 1485 (73.9%) | 1118 (58.7%) | 4391 (68.3%) |
| D and E | 428 (17.0%) | 325 (16.2%) | 392 (20.6%) | 1145 (17.8%) |
| Screen Time |  |  |  |  |
| Mean (SD) | 3.4 (1.9) | 3.9 (2.8) | 3.8 (3.4) | 3.7 (2.6) |
| Median (IQR) | 3 (2-5) | 4 (2-5) | 3 (2-5) | 3 (2-5) |
| SDQ inattention/hyperactivity score<br>(0-10) |  |  |  |  |
| Mean (SD) | 4.9 ( 3.1) | 4.1 (3.1) | 3.5 (3.0) | 4.3 (3.1) |
| Median (IQR) | 5 (2-8)* | 4 (1-7)* | 3 (1-6)* | 4 (2-7) |
| CBCL-ABCL inattention/hyperactivity<br>score (0-16) |  |  |  |  |
| Mean (SD) | 3.7 (3.7) | 3.6 (3.5) | 2.8 (3.2) | 3.4 (3.5) |
| Median (IQR) | 3 (0-6)* | 3 (1-6)* | 2 (0-6)* | 2 (0-5) |

Note. PCPD- Primary Caregiver History of Psychiatric Diagnosis; SDQ - Strengths and Difficulties Questionnaire; CBCL-ABCL - Child Behavior Checklist harmonized with the Adult Behavior Checklist.
\*Differences are significant across time

### Longitudinal associations between inattention/hyperactivity and screen time

The RI-CLPM and the adjusted RI-CLPM presented very good fit indices (RMSEA = 0.016; 95% CI, 0.000, 0.059, CFI = 1.000, SRMR = 0.007; and RMSEA = 0.049; 95% CI, 0.042, 0.055, CFI = 0.0915, SRMR = 0.033 respectively). Results are shown for both models in Table 2 and for the adjusted model in Figure 1. Within-person cross-lagged paths were not significant for both models, indicating no longitudinal relationships between changes in screen time exposure and attention/hyperactivity scores, and vice-versa. However, between-person associations revealed small significant correlations between the Random Intercepts in the SDQ model (β = 0.24; 95% CI, 0.02, 0.45; p = 0.029) that weren’t identified after covariate insertion in the Adjusted SDQ model (β = 0.05; 95% CI, -0.00, 0.45; p = 0.052), suggesting that the underlying factors linking these variables may be explained by confounding variables.

**Figure 1.**
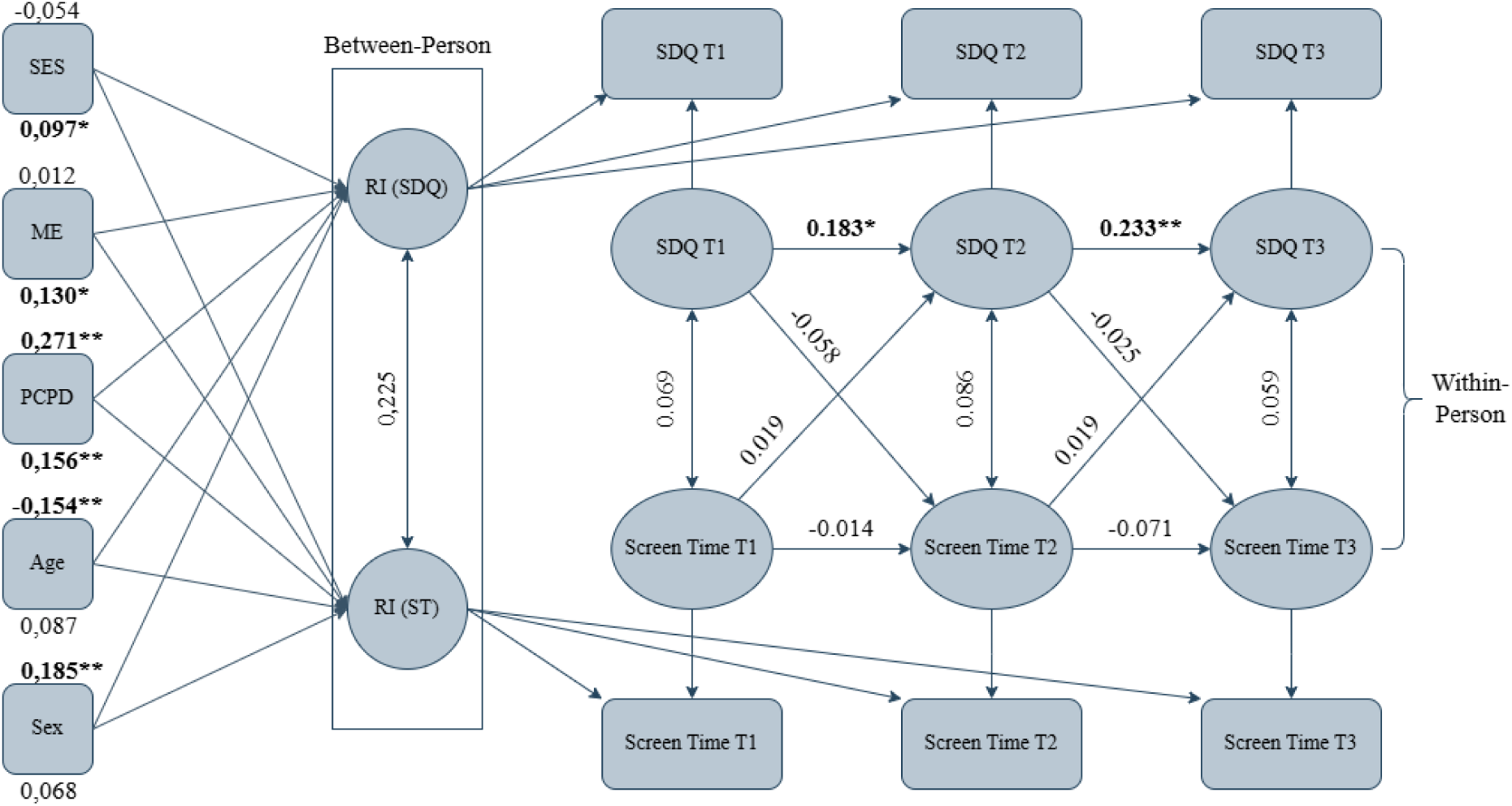
**Longitudinal associations between inattention/hyperactivity and screen time** Note. SE - Socioeconomic class; ME - Maternal education level; PCPD- Primary caregiver history of psychiatric diagnosis; Sex (female as reference); SDQ - Strength and Difficulties Questionnaire; ST - Screen Time; RI - Random Intercept. *p value <0.05; ** p value <0.001

**Table 2.**
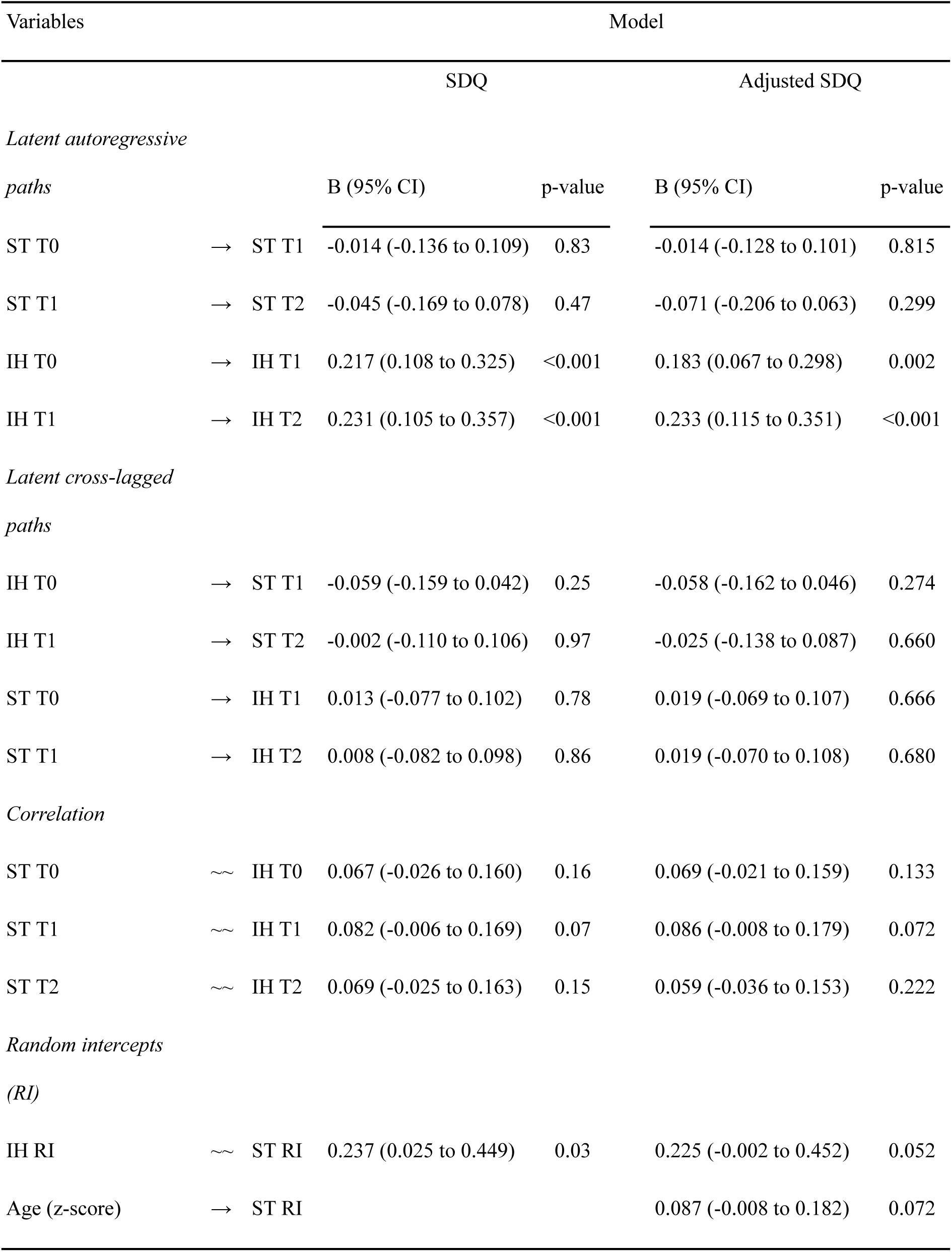
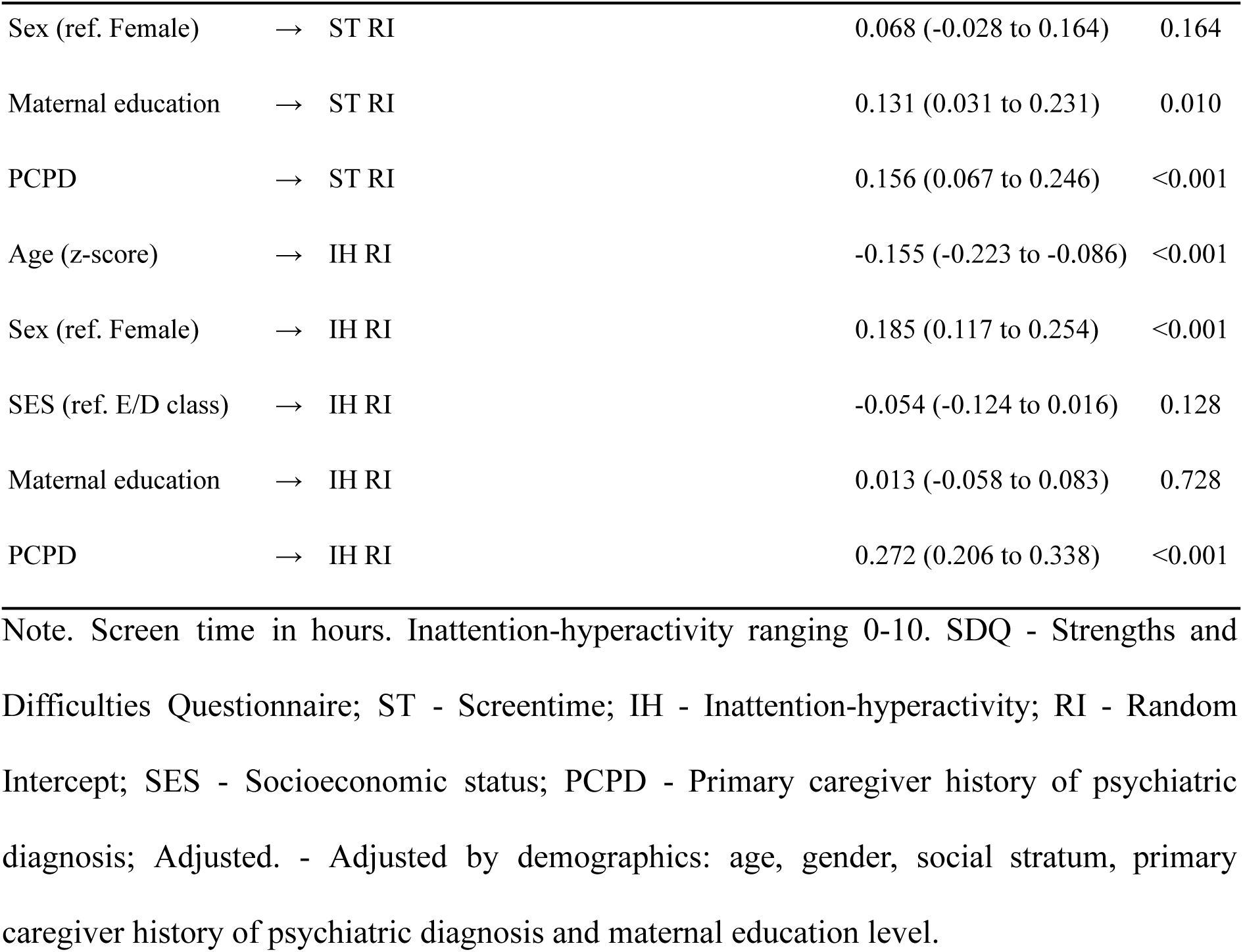
Results of SDQ RI-CPLM Models.

### Covariates relationships with Random Intercepts

Primary caregiver history of psychiatric diagnosis was the only factor significantly associated with both increased general average for higher screen time (β = 0.16; 95% CI, 0.07, 0.25; p < 0.001) and for inattention/hyperactivity scores (β = 0.27; 95% CI, 0.21, 0.34; p < 0.001). Additionally, being male (β = 0.18; 95% CI, 0.12, 0.25; p < 0.001) and older (B = -0.15; 95% CI, -0.22, -0.09; p < 0.001) had small significant association on higher inattention/hyperactivity average. A higher maternal level of education (β = 0.13; 95% CI, 0.03, 0.23; p = 0.010) and higher socioeconomic class (β = 0.010; 95% CI, 0.00, 0.19; p = 0.038) were both linked to a slight increased screen time average (Table 2).

### Sensitivity analysis

The adjusted CBCL-ABCL model showed good RMSEA (0.016; 95% CI, 0-0.059) and SRMR (0.035) fit indices but non optimal CFI value (0.885). Nonetheless, results were qualitatively the same (Table S5) as the previous SDQ models, revealing that results were robust to the questionnaire used (see Supplementary Results for full details).

## Discussion

The present study examined the relationship between general screen time and inattention/hyperactivity symptoms by disentangling between- and within-person associations from late childhood to young adulthood in Brazil, accounting for confounders and questionnaire sensitivity. As hypothesized, our findings suggest that higher than expected screen time use does not increase inattention/hyperactivity symptoms at follow-ups and vice-versa (cross-lagged within-person associations). However, higher average screen time was correlated with higher average inattention/hyperactivity symptoms (between-person association), as we expected, though this link disappeared after adjusting for covariates such as primary caregiver psychiatric history, which was associated with both variables. Also, SDQ and CBCL-ACBCL models were found to have qualitatively the same results, which indicates that the findings are robust to measurement variation.

Our null cross-lagged findings are consistent with studies showing no effect of overall media use (watching TV and playing video games) on inattention/hyperactivity in middle childhood (Beyens et al., 2020), and of video-centric activities (watching TV or videos, and playing video games) from middle childhood to early adolescence (Deng et al., 2024). Similarly, our results align with findings in adolescents showing no cross-lagged effects between social media use and externalizing/inattention symptoms (Beeres et al., 2021), or between social media use intensity and inattention/hyperactivity symptoms (Boer et al., 2020). However, other studies diverge from our findings. Higher externalizing/inattention in early childhood predicted increased general screen time (Neville et al., 2021), inattention/hyperactivity symptoms predicted later violent media use (Beyens et al., 2020), and problematic social media use was linked to later inattention/hyperactivity (Boer et al., 2020). Tiraboschi et al. (2025) reported unidirectional effects between video game use and inattention/hyperactivity symptoms, which reversed across two developmental periods (ages 6–7 and 7–8).

Several factors may explain these discrepancies. First, our study was conducted in a LMIC, whereas all previous research comes from HICs—contexts where socioeconomic and cultural conditions likely influence both media use and symptom expression. In LMICs, for instance, limited resources and teacher shortages may lead schools to rely on screen-based activities like TV as substitutes for instruction. Second, combining inattention/hyperactivity with conduct problems into broad externalizing scores may conflate distinct behavioral processes. Third, specific screen activities (e.g., violent media, media multitasking) may engage different mechanisms than general screen use, which broader measures might overlook. Fourth, we examined the transition from late childhood into adulthood, while prior studies focused only on childhood or adolescence. Finally, earlier studies often used shorter follow-up periods (≤2 years), potentially capturing transient effects that don’t persist long-term. Differences in setting, construct specificity, developmental stage, and follow-up duration likely contribute to the inconsistent findings across studies.

Our between-person findings align with previous RI-CLPM studies (Beeres et al., 2021; Beyens et al., 2020; Boer et al., 2020; Neville et al., 2021), showing significant correlations between screen time and inattention/hyperactivity symptoms in unadjusted models. However, these associations did not persist after adjusting for covariates—contrasting with Deng et al. (2024)—suggesting that much of the observed relationship may be driven by contextual confounders such as socioeconomic status and maternal education. In our LMIC sample, unlike in HIC studies (Deng et al., 2024), higher maternal education and income were associated with greater screen time exposure, likely due to disparities in device access—only 39% of Brazilian households owned a computer or laptop in 2023 (IBGE, 2023), compared to 80.5% in the U.S. in 2021 (ACSR, 2024). Notably, caregiver psychiatric history was the only covariate linked to both increased screen time and higher inattention/hyperactivity symptoms, possibly reflecting both genetic vulnerability and reduced parental capacity to regulate screen use in households affected by mental health challenges.

Regarding limitations, firstly our sample did not include toddlers, which may have constrained our understanding of screen time’s impact as emerging evidence seems to link early screen time exposure to emotion regulation difficulties (Fitzpatrick et al., 2024). Second, screen time was assessed solely through parent reports, which may lack agreement with self-reports (Borodovsky, Squeglia, Mewton & Marsch, 2024), and do not match the precision of the golden standard ecological momentary assessment (Shiffman, Stone & Hufford, 2008). Third, our broad screen time construct doesn’t distinguish between different media types (e.g., passive TV viewing vs. interactive gaming) or context and motive (e.g., educational vs. recreational), which may yield distinct effects on attention, and it lacks cell phone and tablet screen times. Also, our long timed intervals (3-5 years) between assessments may obscure shorter-term effects. However, this does not invalidate present findings as our study demonstrates no long-lasting associations using a wider time frame.

## Conclusions

Our study suggests no relationship between changes in screen time exposure and inattention/hyperactivity symptomatology in late childhood through adulthood. Between-person effects were identified in the unadjusted SDQ model, but when comparing with adjusted models the average effect was found to be explained by covariates. Primary caregiver history of psychiatric diagnosis influenced both general inattention/hyperactivity symptomatology and general screen time exposition averages. Future research should employ RI-CLPM designs with covariate adjustment, considering different media types, context and motive of use, and different inattention/hyperactivity questionnaires to check if findings are consistent across different measures.

## Supporting information

Supplementary Methods Results Tables

Code and Statistical Analysis Syntax

## Supporting information Supplementary Methods

Design of confirmatory factor analysis.

## Code and Statistical Analysis Syntax

Code and Statistical Analysis Syntax used to model and apply CFA, RI-CLPM, Shapiro-Wilk and Wilcoxon signed-rank tests.

## Supplementary Results

Results from CFA, Shapiro-Wilk and Wilcoxon signed-rank tests for inattention/hyperactivity symptoms, and sensitivity analysis.

**Table S1.** Standard SDQ 4F and CBCL-ABCL CFA fits.

**Table S2.** SDQ 4 factors structure.

**Table S3.** CBCL-ABCL eight-syndrome model.

**Table S4.** Shapiro-Wilk and Wilcoxon signed-rank tests for inattention/hyperactivity symptoms.

**Table S5.** Results of CBCL-ABCL RI-CPLM Model.

## Data Availability

Data dictionary is available at https://osf.io/ktz5h/wiki/Data%20Dictionaries/ and https://osf.io/w3jr4 to direct download. Individual-level data is available upon request to the Brazilian High-Risk Cohort Study research committee, by following the instructions and filling the research form available at https://osf.io/ktz5h/wiki/home/.

## Acknowledgements

This work is supported by the National Institute of Developmental Psychiatry for Children and Adolescents, a science and technology institute funded by Conselho Nacional de Desenvolvimento Científico e Tecnológico (CNPq; National Council for Scientific and Technological Development; grant numbers 573974/2008-0 and 465550/2014-2) and Fundação de Amparo à Pesquisa do Estado de São Paulo (FAPESP; Research Support Foundation of the State of São Paulo; grant number 2008/57896-8, 2014/50917-0 and 2021/05332-8) to the National Institute of Development Psychiatric for Children and Adolescent (INPD). This work was also funded by the European Research Council under the European Union’s Seventh Framework Programme (FP7/2007-2013)/ERC grant agreement no 337673.

Ricardo Kaciava Bombardelli was supported by FIPE/UFSM 2025 scholarship, granted to prof. Mauricio Scopel Hoffmann. Dr. Mauricio Scopel Hoffmann is supported by the United States National Institutes of Health grant R01MH120482 under his post-doctoral fellowship at UFRGS.

## Ethical information

This study was approved by the National Research Ethics Commission (Comissão Nacional de Ética em Pesquisa) under the approval number 2.448.062 (CAAE 74563817.7.1001.5327), University of São Paulo, and Federal University of Rio Grande do Sul ethics committees. Written informed consent was obtained from parents and participants who were able to read, write, and clearly understand the written consent. From those who were illiterate, the consent was read, doubts were explained, and verbal agreement was obtained. None were excluded due to parental illiteracy (Salum et al., 2015).

## Key points

● Screen time has been linked to inattention/hyperactivity, but findings on directionality remain mixed.
● It is unclear whether this association reflects within-person effects over time, particularly in LMIC settings.
● We applied random intercept cross-lagged panel models to data from a Brazilian cohort (mean ages 10–18).
● Changes in screen time did not predict changes in inattention/hyperactivity, and between-person associations disappeared after adjusting for covariates (e.g., caregiver psychiatric history).

Screen time may not be a causal driver of inattention/hyperactivity and interventions should consider broader contextual factors.

