## Supplementary Methods Results Tables for "Testing Bidirectional Associations Between Screen Time and Inattention/Hyperactivity Symptoms From Childhood to Adulthood in a Brazilian Cohort"

### Confirmatory factor analysis for CBCL-ABCL and SDQ

The CBCL-ABCL model is composed of eight dimensions, namely anxious-depressed (12 items), withdrawal-depressed (8 items), somatic complaints (9 items), rule-breaking (10 items), aggressive behavior (16 items), social problems (9 items), thought problems (11 items) and hyperactivity/inattention dimension (8 items), which was used for the purpose of the sensitivity analysis. All CFA considered the subject as a cluster to estimate a model, and factor scores, considering the longitudinal characteristic of the study. The analysis was implemented with Weighted Least Square Mean and Variance (WLSMV) as the estimation method. Model goodness of fit was evaluated using root-mean-square error of approximation (RMSEA), comparative fit index (CFI), Tucker-Lewis index (TLI) and Standardized Root Mean Square Residual (SRMR). RMSEA lower than 0.060 and CFI or TLI values higher than 0.950 indicate a good-to-excellent model. SRMR lower or equal than 0.100 indicate adequate fit, and lower than 0.060 in combination with previous indices indicate good fit (Hu & Bentler, 1999).

In order to assess the reliability of the factors, we considered Lucke's omega (ω; Lucke, 2005), a model-based reliability estimate, being analogous to alpha coefficient but appropriate for congeneric tests (varying factor loadings); Values of ω may vary between 0 and 1, where higher scores indicate greater reliability; a value of 1 indicates that the instrument's sum score measures the target construct with perfect accuracy. We also estimated factor determinacy (FD, the correlation between the factor scores and the estimated factor, indicating that factor scores should be used if at least 0.9) and H index (a measure of construct replicability that quantifies how well each latent factor is represented by the items loading on it, with $H > 0.7$ representing a well-defined latent variable according to Hancock

& Mueller (2001). Longitudinal measurement invariance for the SDQ model was implemented by specifying the waves of data collection in the grouping function and specifying "model = configural scalar" in Mplus. Invariance decision was based on ΔCFI < 0.010 supplemented by ΔRMSEA < 0.015 or ΔSRMR < 0.010 (Chen, 2007). CFA was performed using Mplus version 8.6 (Muthén & Muthén, 2017) and implemented in RStudio version 2024.04.2+764 and R version 4.4.1 using the *MplusAutomation* package (Hallquist & Wiley, 2018), which was also used to extract factor scores generated in Mplus. All bifactor reliability indices were calculated using the *BifactorIndicesCalculator* package in R (Dueber, 2017).

CFA was performed using Mplus version 8.6 (Muthén & Muthén, 2017) and implemented in RStudio version 2024.04.2+764 and R version 4.4.1 using the *MplusAutomation* package (Hallquist & Wiley, 2018), which was also used to extract factor scores generated in Mplus. All bifactor reliability indices were calculated using the *BifactorIndicesCalculator* package in R (Dueber, 2017).

### Supplementary Results

#### Confirmatory factor analysis

The model fit indices are summarized in Table S1. Overall, all models demonstrated acceptable fit, with the exception of the SDQ four-factor scalar model, which had suboptimal CFI values. Both CBCL-ACBCL models exhibited good RMSEA fit, while the SDQ models showed acceptable RMSEA values. Additionally, all models achieved adequate SRMR indices. In terms of invariance testing, no significant differences were observed between the configural and scalar models for either questionnaire.

The standardized factor loadings of the SDQ 4-factor structural model, as presented in Table S2, ranged from 0.32 to 0.81 across all items, reflecting that relationships between observed items and their respective latent factors varied from small to strong. Within each factor, specific items demonstrated the strongest and weakest loadings: for the Emotional factor, "Unhappy" had the highest loading (0.78), while "Worried" had the lowest (0.44); in the Hyperactive factor, "Distracted" showed the strongest loading (0.81), and "Think before acting" the weakest (0.62); for the Conduct Problems factor, "Loses temper" had the highest loading (0.78), and "Well-behaved" the lowest (0.52); and in the Peer-relationship problems factor, "Bullied" exhibited the strongest loading (0.73), while "Solitary" had the weakest (0.39). Regarding correlation between factors, the strongest relationship was found to be between Hyperactivity and Conduct Problems factors (0.81), while the weakest relationship was among Hyperactivity and Peer-relationship problems factors (0.60).

In terms of reliability indices, most factors demonstrated good Factor Determinacy (FD) and H-index, indicating that most factor scores are well-defined, reliable, and appropriate for use in further analyses (Table S2). However, the Peer-relationship problems factor showed borderline acceptable Factor Determinacy (0.88) and a poor H-index (0.697). Regarding Lucke's Omega (ω), the Emotional (0.68) and Peer-relationship problems (0.54) factors exhibited poor reliability values, while the remaining factors displayed acceptable reliability.

The standardized factor loadings for the CBCL-ABCL structural model, detailed in Table S3, spanned from 0.28 to 0.94 across all items, indicating a spectrum of relationships from small to strong between the observed items and their corresponding latent factors. Within each factor, certain items stood out for their strong or weak loadings: in the Anxious-depressed factor, "Worthless" and "Nervous" had the highest loadings (0.83), whereas "Perfect" had the lowest (0.28); for the Withdrawn-depressed factor, "Sad" showed the strongest loading (0.90), and "Shy" the weakest (0.52); in the Somatic factor, "Tired" had the highest loading (0.93), while "Skin problems" had the lowest (0.50); and within the Rule-breaking factor, "Bad friends" exhibited the strongest loading (0.87), and "Prefers older" the weakest (0.48). Similarly, in the Aggressive factor, "Sulks" had the highest loading (0.83), and "Teases" the lowest (0.58); for the Social problems factor, "Feels persecuted" showed the strongest loading (0.72), while "Speech problem" had the weakest (0.40); in the Thought problems factor, "Strange behavior" had the highest loading (0.80), and "Picks skin" the lowest (0.51); and within the Attention problems factor, "Confused" exhibited the strongest loading (0.84), while "Poor school work" had the weakest (0.66). Additionally, correlations between factors revealed the strongest relationship between the Anxious-depressed and Social problems factors (0.94), and the weakest between the Rule-breaking and Somatic factors (0.49)

Regarding reliability indices, all factors demonstrated acceptable Factor Determinacy (FD) and H-index values, indicating that the factor scores, including Attention problems, are well-defined, reliable, replicable and suitable for use in further analyses. (Table S3). As for Lucke's Omega (ω), all factors exhibited great reliability.

**Shapiro-Wilk and Wilcoxon signed-rank tests**

Results are shown in Table S4. Shapiro-Wilk tests found no normally distributed inattention/hyperactivity scores at any time point in either SDQ or CBCL-ACBCL questionnaire. For SDQ inattention/hyperactivity scores Wilcoxon signed-rank tests indicated a significant decline in median scores from 5 (IQR: 2-8) to 4 (IQR: 1-7) between baseline and wave 1 ($V = 936216$, $p < 0.001$) and to 3 (IQR: 1-6) between wave 1 and wave 2 ($V = 499011$, $p < 0.001$). For CBCL-ABCL inattention/hyperactivity scores Wilcoxon signed-rank tests showed a significant difference in scores between baseline and wave 1 (V = 703345, p =

0.010) and a significant decrease from 3 (IQR: 1-6) to 2 (IQR: 0-6) between wave 1 and wave 2 ($V = 363501$, $p < 0.001$). While the median remained stable from the baseline and first wave, the mean declined slightly from 3.72 to 3.55, suggesting a small overall reduction in scores.

**CBCL-ACBCL Random Intercepts Cross Lagged Panel Model**

Results are shown in Table S5. Within-person cross-lagged paths were not significant, indicating no longitudinal relationships between changes in screen time exposure and attention/hyperactivity scores, and vice-versa. Additionally, between-person associations also revealed non-significant correlations between the Random Intercepts, suggesting the absence of shared underlying factors linking these variables or that they are explained by covariates inserted in this model which aligns with the adjusted SDQ model. Autoregressive paths for attention/hyperactivity factor scores demonstrated stability over time: from baseline to wave 1 ($\beta = 0.14$; 95% CI, 0.01, 0.27; $p = 0.041$), and from wave 1 to wave 2 ($\beta = 0.16$; 95% CI, 0.03, 0.30; $p = 0.019$). No significant autoregressive paths for screen time were found in these models.

When examining the relationships between Random Intercepts and covariates, maternal psychiatric diagnosis was linked to tendencies for both higher inattention/hyperactivity scores ($\beta = 0.33$; 95% CI, 0.26, 0.40; $p < 0.001$) and screen time ($\beta = 0.16$; 95% CI, 0.07, 0.25; $p = 0.001$), while higher socioeconomic status was associated with tendencies for lower inattention/hyperactivity scores ($\beta = -0.06$; 95% CI, -0.13, -0.00; $p = 0.048$) and higher screen time ($\beta = 0.09$; 95% CI, 0.00, 0.19; $p = 0.045$). Additionally, being male was associated with the tendency for higher inattention/hyperactivity scores ($\beta = 0.17$; 95% CI, 0.11, 0.24; $p < 0.001$), while higher maternal education level had a significant association with the tendency for higher screen time ($\beta$ = 0.13; 95% CI, 0.03, 0.23; $p$ = 0.011).

### Tables and Figures

**Table S1**

*Standard SDQ 4F and CBCL-ABCL CFA fits*

| | CFI | RMSEA | SMRM | ΔCFI | ΔRMSEA | ΔSMRM | Decision |
|---|---|---|---|---|---|---|---|
| SDQ 4 Factors Configural | 0.902 | 0.069 | 0.071 | | | | |
| SDQ 4 Factors Scalar | 0.888 | 0.07 | 0.075 | 0.014 | 0.001 | 0.004 | Accepted |
| CBCL-ABCL Configural | 0.929 | 0.032 | 0.075 | | | | |
| CBCL-ABCL Scalar | 0.932 | 0.031 | 0.076 | 0.003 | 0.001 | 0.001 | Accepted |

Note. SDQ - Strengths and Difficulties Questionnaire; CBCL-ABCL - Child Behavior Checklist harmonized with the Adult Behavior Checklist; CFI - Comparative Fit Index; RMSEA - Root Mean Square Error of Approximation; SRMR - Standardized Root Mean Square Residual.

**Table S2**

*SDQ 4 factors structure*

| | Factor Reliability | Factors and factor loadings | | | |
|---|---|---|---|---|---|
| | | Emot. | Hyper. | CP | PRP |
| | ω | 0.684 | 0.789 | 0.717 | 0.545 |
| | H index | 0.801 | 0.869 | 0.817 | 0.697 |
| | FD | 0.913 | 0.943 | 0.931 | 0.880 |
| Content | Item | | | | |
| Worried | PSDQ_8 | 0.437 | | | |
| Unhappy | PSDQ_13 | 0.782 | | | |
| Nervous | PSDQ_16 | 0.682 | | | |
| Many fears | PSDQ_24 | 0.676 | | | |
| Somatic complaints | PSDQ_3 | 0.577 | | | |
| Overactive | PSDQ_2 | | 0.760 | | |
| Fidgety | PSDQ_10 | | 0.788 | | |
| Distracted | PSDQ_15 | | 0.809 | | |
| Think before acting | PSDQ_21 | | 0.621 | | |
| Good attention | PSDQ_25 | | 0.721 | | |
| Loses temper | PSDQ_5 | | | 0.778 | |
| Well-behaved | PSDQ_7 | | | 0.519 | |
| Fight others | PSDQ_12 | | | 0.736 | |
| Lies or cheats | PSDQ_18 | | | 0.699 | |
| Steals | PSDQ_22 | | | 0.545 | |
| Solitary | PSDQ_6 | | | | 0.390 |

(continues)

**Table S2** continued

| | | | | | |
|---|---|---|---|---|---|
| Has a friend | PSDQ_11 | | | | 0.504 |
| Liked by others | PSDQ_14 | | | | 0.585 |
| Bullied | PSDQ_19 | | | | 0.732 |
| Prefer adults | PSDQ_23 | | | | 0.316 |
| Correlation between factors | | | | | |
| | | Hyper. | CP | PRP | |
| Emot. | | 0.656 | 0.669 | 0.682 | |
| Hyper. | | | 0.811 | 0.603 | |
| CP | | | | 0.732 | |

Note. ω - Lucke's Omega; FD - Factor Determinacy; Emot. - Emotional; Hyper.- Hyperactivity; CP - Conduct Problems; PRP - Peer-relationship Problems.

**Table S3**

*CBCL eight-syndrome bifactor model*

| | | | | | | | | | |
|---|---|---|---|---|---|---|---|---|---|
| | | Factors and factor loadings | | | | | | | |
| | Factor Reliability | AD | WD | Som. | RB | Agg. | SP | TP | AP |
| | ω | 0.805 | 0.845 | 0.809 | 0.791 | 0.909 | 0.779 | 0.823 | 0.849 |
| | H index | 0.921 | 0.929 | 0.93 | 0.898 | 0.956 | 0.864 | 0.913 | 0.92 |
| | FD | 0.975 | 0.967 | 0.967 | 0.965 | 0.983 | 0.979 | 0.966 | 0.969 |
| Content | Item | | | | | | | | |
| Cries | CBCL_14 | 0.676 | | | | | | | |
| Fears | CBCL_29 | 0.417 | | | | | | | |
| Fears do bad | CBCL_31 | 0.371 | | | | | | | |
| Perfect | CBCL_32 | 0.283 | | | | | | | |
| Unloved | CBCL_33 | 0.823 | | | | | | | |
| Worthless | CBCL_35 | 0.826 | | | | | | | |
| Nervous | CBCL_45 | 0.826 | | | | | | | |
| Fearful | CBCL_50 | 0.696 | | | | | | | |
| Feels too guilty | CBCL_52 | 0.703 | | | | | | | |
| Self-conscious | CBCL_71 | 0.621 | | | | | | | |
| Talks about suicide | CBCL_91 | 0.700 | | | | | | | |
| Worries | CBCL_112 | 0.499 | | | | | | | |
| Little they enjoy | CBCL_5 | | 0.832 | | | | | | |
| Prefers alone | CBCL_42 | | 0.715 | | | | | | |
| Won’t talk | CBCL_65 | | 0.772 | | | | | | |
| Secretive | CBCL_69 | | 0.663 | | | | | | |

(continues)

**Table S3** continued

| | | | | |
|---|---|---|---|---|
| Shy | CBCL_75 | 0.521 | | |
| Lacks energy | CBCL_102 | 0.737 | | |
| Sad | CBCL_103 | 0.897 | | |
| Withdrawn | CBCL_111 | 0.801 | | |
| Dizzy | CBCL_51 | | 0.755 | |
| Tired | CBCL_54 | | 0.933 | |
| Aches | CBCL_56A | | 0.719 | |
| Headaches | CBCL_56B | | 0.668 | |
| Nausea | CBCL_56C | | 0.734 | |
| Eye problems | CBCL_56D | | 0.495 | |
| Skin problems | CBCL_56E | | 0.370 | |
| Stomach | CBCL_56F | | 0.687 | |
| Vomit | CBCL_56G | | 0.692 | |
| No guilt | CBCL_26 | | | 0.672 |
| Breaks rules at home | CBCL_28 | | | 0.866 |
| Bad friends | CBCL_39 | | | 0.654 |
| Lies or cheats | CBCL_43 | | | 0.753 |
| Prefers older | CBCL_63 | | | 0.480 |
| Steals outside home | CBCL_82 | | | 0.560 |
| Swears | CBCL_90 | | | 0.710 |
| Thinks about sex | CBCL_96 | | | 0.502 |
| Truants | CBCL_101 | | | 0.590 |
| Use drugs | CBCL_105 | | | 0.481 |

(continues)

**Table S3** continued

| | | | |
|---|---|---|---|
| Argues | CBCL_3 | 0.689 | |
| Mean | CBCL_16 | 0.681 | |
| Demands a lot of attention | CBCL_19 | 0.733 | |
| Destroys own things | CBCL_20 | 0.742 | |
| Destroys other | CBCL_21 | 0.777 | |
| Disobedient at home | CBCL_22 | 0.774 | |
| Fights | CBCL_37 | 0.723 | |
| Attacks | CBCL_57 | 0.723 | |
| Screams | CBCL_68 | 0.736 | |
| Stubborn | CBCL_86 | 0.833 | |
| Mood changes | CBCL_87 | 0.834 | |
| Sulks | CBCL_88 | 0.835 | |
| Teases | CBCL_94 | 0.583 | |
| Temper | CBCL_95 | 0.814 | |
| Threatens | CBCL_97 | 0.744 | |
| Loud | CBCL_104 | 0.663 | |
| Too dependent | CBCL_11 | | 0.569 |
| Lonely | CBCL_12 | | 0.689 |
| Doesn’t get along | CBCL_25 | | 0.627 |
| Jealous | CBCL_27 | | 0.685 |
| Feels persecuted | CBCL_34 | | 0.720 |
| Gets hurt a lot | CBCL_36 | | 0.606 |
| Not liked by peers | CBCL_48 | | 0.645 |

(continues)

**Table S3** continued

| | | | | |
|---|---|---|---|---|
| Clumsy | CBCL_62 | 0.686 | | |
| Speech problem | CBCL_79 | 0.407 | | |
| Mind off | CBCL_9 | | 0.759 | |
| Harms self | CBCL_18 | | 0.644 | |
| Hears things | CBCL_40 | | 0.714 | |
| Twitches | CBCL_46 | | 0.607 | |
| Picks skin | CBCL_58 | | 0.511 | |
| Repeats acts | CBCL_66 | | 0.733 | |
| Sees things | CBCL_70 | | 0.703 | |
| Stores up | CBCL_83 | | 0.609 | |
| Strange behavior | CBCL_84 | | 0.799 | |
| Strange ideas | CBCL_85 | | 0.748 | |
| Sleep problems | CBCL_100 | | 0.622 | |
| Fails to finish things | CBCL_4 | | | 0.760 |
| Can't concentrate | CBCL_8 | | | 0.753 |
| Can't sit still | CBCL_10 | | | 0.707 |
| Confused | CBCL_13 | | | 0.839 |
| Daydreams | CBCL_17 | | | 0.748 |
| Impulsive | CBCL_41 | | | 0.831 |
| Poor school work | CBCL_61 | | | 0.658 |
| Stares | CBCL_80 | | | 0.730 |
| Correlation between factors | | | | |

(continues)

**Table S3** continued

| | | | | | | | |
|---|---|---|---|---|---|---|---|
| AD | 0.791 | 0.749 | 0.638 | 0.775 | 0.941 | 0.855 | 0.746 |
| WD | | 0.626 | 0.548 | 0.571 | 0.719 | 0.702 | 0.635 |
| Som. | | | 0.487 | 0.577 | 0.672 | 0.679 | 0.578 |
| RB | | | | 0.908 | 0.798 | 0.696 | 0.819 |
| Agg. | | | | | 0.881 | 0.766 | 0.847 |
| SP | | | | | | 0.878 | 0.891 |
| TP | | | | | | | 0.795 |

Note: ω - Lucke’s Omega; FD - Factor Determinacy; AD - Anxious-depressed; WD - Withdraw-depressed; Som. - Somatic; RB - Rule breaking; Agg. - Aggressive; SP - Social Problems; TP - Thought problems; AP - Attention problems.

**Table S4**

*Shapiro-Wilk and Wilcoxon signed-rank tests for ADHD scores*

| Shapiro-Wilk | | | Wilcoxon signed-rank | | |
|---|---|---|---|---|---|
| | W | p-value | | V | p-value |
| SDQ Baseline | 0.95 | < 0.001 | SDQ | | |
| SDQ W1 | 0.93 | < 0.001 | Baseline - W1 | 936216 | < 0.001 |
| SDQ W2 | 0.91 | < 0.001 | W1 - W2 | 499011 | < 0.001 |
| CBCL-ABCL Baseline | 0.88 | < 0.001 | CBCL-ABCL | | |
| CBCL-ABCL W1 | 0.88 | < 0.001 | Baseline - W1 | 703345 | 0.010 |
| CBCL-ABCL W2 | 0.83 | < 0.001 | W1 - W2 | 363501 | < 0.001 |

Note. SDQ - Strengths and Difficulties Questionnaire; CBCL-ABCL - Child Behavior Checklist harmonized with the Adult Behavior Checklist; V- Shapiro-Wilk V statistic; W - Wilcoxon signed-rank W statistic

**Table S5**

*Results of CBCL-ABCL RI-CPLM Model*

| Variables | | | Model | |
|---|---|---|---|---|
| | | | Adjusted CBCL-ABCL | |
| *Latent autoregressive paths* | | | B (95% CI) | p-value |
| ST T0 | → | ST T1 | -0.014 (-0.128 to 0.101) | 0.814 |
| ST T1 | → | ST T2 | -0.081 (-0.214 to 0.053) | 0.237 |
| IH T2 | → | IH T1 | 0.138 (0.006 to 0.270) | 0.041 |
| IH T1 | → | IH T2 | 0.162 (0.026 to 0.298) | 0.019 |
| *Latent cross-lagged paths* | | | | |
| IH T0 | → | ST T1 | 0.007 (-0.089 to 0.103) | 0.888 |
| IH T1 | → | ST T2 | 0.071 (-0.035 to 0.177) | 0.191 |
| ST T0 | → | IH T1 | 0.086 (-0.001 to 0.172) | 0.052 |
| ST T1 | → | IH T2 | 0.035 (-0.054 to 0.124) | 0.442 |
| *Correlation* | | | | |
| ST T0 | ~~ | IH T0 | 0.086 (0.002 to 0.171) | 0.044 |
| ST T1 | ~~ | IH T1 | 0.129 (0.035 to 0.223) | 0.007 |
| ST T2 | ~~ | IH T2 | 0.099 (-0.002 to 0.201) | 0.055 |
| *Random intercepts (RI)* | | | | |
| IH RI | ~~ | ST RI | -0.039 (-0.271 to 0.192) | 0.739 |
| Age (z-score) | → | ST RI | 0.090 (-0.006 to 0.186) | 0.065 |
| Sex (ref. Female) | → | ST RI | 0.070 (-0.027 to 0.166) | 0.156 |
| SES (ref. E/D class) | → | ST RI | 0.095 (0.002 to 0.188) | 0.045 |
| Maternal education | → | ST RI | 0.132 (0.031 to 0.233) | 0.011 |

(continues)

**Table S5** continued

| | | | | |
|---|---|---|---|---|
| PCPD | → | ST RI | 0.158 (0.067 to 0.248) | <0.001 |
| Age (z-score) | → | IH RI | -0.065 (-0.131 to -0.000) | 0.050 |
| Sex (ref. Female) | → | IH RI | 0.175 (0.106 to 0.244) | <0.001 |
| SES (ref. E/D class) | → | IH RI | -0.065 (-0.129 to -0.000) | 0.048 |
| Maternal education | → | IH RI | -0.002 (-0.071 to 0.066) | 0.946 |
| PCPD | → | IH RI | 0.331 (0.258 to 0.404) | <0.001 |

Note: Screen time in hours. Inattention-hyperactitivy scores rainging 0-16: CBCL-ABCL - Child Behavior Checklist harmonized with the Adult Behavior Checklist; ST - Screentime; IH - Inattention-hiperactivity; RI - Random Intercept; SES - Socioeconomic status; PCD - Primary caregiver history of psychiatric diagnosis; Adjusted. - Adjusted by demographics: age, gender, social strat, primary caregiver history of psychiatric diagnosis and maternal education level.

,
