## Supplementary material for "Testing Bidirectional Associations Between Screen Time and Inattention/Hyperactivity Symptoms From Childhood to Adulthood in a Brazilian Cohort": Code and Statistical Analysis Syntax: Code_and_Statistical_Analysis_Syntax.html

Screen time (TV, computer or videogame) and Inattention/Hyperactivity - data analysis


### Screen time (TV, computer or videogame) and Inattention/Hyperactivity - data analysis

###### 15 August 2024

### CFA for SDQ and CBCL models

```
#Data set for MPlus
mydata_long_mplus <- mydata_long %>% dplyr::select(ident, wave, CBCL_1:sSDQ_20)
mydata_long_mplus$id_random <- mydata_long_mplus$ident+100000
mydata_long_mplus[is.na(mydata_long_mplus)] <- 9999 # MISSING ARE ALL (9999);

names(mydata_long_mplus) <- NULL # exclude header
rio::export(mydata_long_mplus, "ADHD_Screentime_mplus_models.txt") #export data to be used in all models

##Models are run in Mplus
#runModels("ADHD_Screentime_SDQ_4F_Model.inp") 
#runModels("ADHD_Screentime_CBCL_ABCL_Model.inp") 
#runModels("ADHD_Screentime_SDQ_4F_invariance_testing.inp") 
#runModels("ADHD_Screentime_CBCL_ABCL_invariance_testing.inp") 

#Extracting models from Mplus
results_SDQ_model <- readModels("ADHD_Screentime_SDQ_4F_Model.out") 
results_CBCL_model <- readModels("ADHD_Screentime_CBCL_ABCL_Model.out") 
results_SDQ_invariance_testing <- readModels("ADHD_Screentime_SDQ_4F_invariance_testing.out") 
results_CBCL_ABCL_invariance_testing <- readModels("ADHD_Screentime_CBCL_ABCL_invariance_testing.out") 

#Extracting model fit from all output files (.out) #
ModelFit_SDQ_model <- results_SDQ_model[["summaries"]]
ModelFit_CBCL_model <- results_CBCL_model[["summaries"]]
ModelFit_SDQ_invariance_testing <- results_SDQ_invariance_testing[["summaries"]]
ModelFit_CBCL_ABCL_invariance_testing <- results_CBCL_ABCL_invariance_testing[["summaries"]]

Table_ModelFit_SDQ_model <- ModelFit_SDQ_model %>% dplyr::select(Title, RMSEA_Estimate, RMSEA_90CI_LB, RMSEA_90CI_UB, CFI, TLI, SRMR)
Table_ModelFit_SDQ_model <- Table_ModelFit_SDQ_model %>% dplyr::rename(Model = Title) %>% dplyr::rename(RMSEA = RMSEA_Estimate) %>% dplyr::rename(RMSEA_LB = RMSEA_90CI_LB) %>% dplyr::rename(RMSEA_UB = RMSEA_90CI_UB)

Table_ModelFit_CBCL_model <- ModelFit_CBCL_model %>% dplyr::select(Title, RMSEA_Estimate, RMSEA_90CI_LB, RMSEA_90CI_UB, CFI, TLI, SRMR)
Table_ModelFit_CBCL_model <- Table_ModelFit_CBCL_model %>% dplyr::rename(Model = Title) %>% dplyr::rename(RMSEA = RMSEA_Estimate) %>% dplyr::rename(RMSEA_LB = RMSEA_90CI_LB) %>% dplyr::rename(RMSEA_UB = RMSEA_90CI_UB)
Table_ModelFit_SDQ_CBCL_models <- rbind(Table_ModelFit_SDQ_model, Table_ModelFit_CBCL_model)

Table_ModelFit_SDQ_invariance_testing <- ModelFit_SDQ_invariance_testing %>% dplyr::select(Title, RMSEA_Estimate, RMSEA_90CI_LB, RMSEA_90CI_UB, CFI, TLI, SRMR)
Table_ModelFit_SDQ_invariance_testing <- Table_ModelFit_SDQ_invariance_testing %>% dplyr::rename(Model = Title) %>% dplyr::rename(RMSEA = RMSEA_Estimate) %>% dplyr::rename(RMSEA_LB = RMSEA_90CI_LB) %>% dplyr::rename(RMSEA_UB = RMSEA_90CI_UB)
Table_ModelFit_SDQ_CBCL_models <- rbind(Table_ModelFit_SDQ_CBCL_models, Table_ModelFit_SDQ_invariance_testing)

Table_ModelFit_CBCL_ABCL_invariance_testing <- ModelFit_CBCL_ABCL_invariance_testing %>% dplyr::select(Title, RMSEA_Estimate, RMSEA_90CI_LB, RMSEA_90CI_UB, CFI, TLI, SRMR)
Table_ModelFit_CBCL_ABCL_invariance_testing <- Table_ModelFit_CBCL_ABCL_invariance_testing %>% dplyr::rename(Model = Title) %>% dplyr::rename(RMSEA = RMSEA_Estimate) %>% dplyr::rename(RMSEA_LB = RMSEA_90CI_LB) %>% dplyr::rename(RMSEA_UB = RMSEA_90CI_UB)
Table_ModelFit_SDQ_CBCL_models <- rbind(Table_ModelFit_SDQ_CBCL_models, Table_ModelFit_CBCL_ABCL_invariance_testing)

write.xlsx(Table_ModelFit_SDQ_CBCL_models, 
           file = "ADHD_Screentime_Results.xlsx", 
           sheetName = "Model Fit", 
           overwrite = TRUE)

#Exctracting reliability
results_SDQ_indices <- BifactorIndicesCalculator::bifactorIndicesMplus(Lambda = "ADHD_Screentime_SDQ_4F_Model.out")
results_SDQ_FactorLevelIndices <- results_SDQ_indices[["FactorLevelIndices"]]

results_CBCL_indices <- BifactorIndicesCalculator::bifactorIndicesMplus(Lambda = "ADHD_Screentime_CBCL_ABCL_Model.out")
results_CBCL_FactorLevelIndices <- results_CBCL_indices[["FactorLevelIndices"]]

results_SDQ_CBCL_FactorLevelIndices <- rbind(results_SDQ_FactorLevelIndices, results_CBCL_FactorLevelIndices)
results_SDQ_CBCL_FactorLevelIndices <- as.data.frame(results_SDQ_CBCL_FactorLevelIndices)
results_SDQ_CBCL_FactorLevelIndices <- rownames_to_column(results_SDQ_CBCL_FactorLevelIndices, var = "Factors")

wb <- loadWorkbook("ADHD_Screentime_Results.xlsx")
addWorksheet(wb, sheetName = "SDQ CBCL factor Reliability")
writeData(wb, sheet = "SDQ CBCL factor Reliability", x = results_SDQ_CBCL_FactorLevelIndices)
saveWorkbook(wb, file = "ADHD_Screentime_Results.xlsx", overwrite = TRUE)

##Exctracting factor loadings
#SDQ structure
Table_FL_SDQ <- results_SDQ_model[["parameters"]][["stdyx.standardized"]] 
Table_FL_SDQ <- Table_FL_SDQ %>% dplyr::select(paramHeader, param, est, se, pval)
Table_FL_SDQ <- Table_FL_SDQ %>% dplyr::rename(Factor = paramHeader) %>% dplyr::rename(Item = param) %>% dplyr::rename(Loading = est) %>% dplyr::rename(SE = se) %>% dplyr::rename("P-value" = pval)
Table_FL_SDQ <- Table_FL_SDQ[Table_FL_SDQ$Factor != "Thresholds", ]
Table_FL_SDQ <- Table_FL_SDQ[Table_FL_SDQ$Factor != "Variances", ]

wb <- loadWorkbook("ADHD_Screentime_Results.xlsx")
addWorksheet(wb, sheetName = "SDQ 4F structure")
writeData(wb, sheet = "SDQ 4F structure", x = Table_FL_SDQ)
saveWorkbook(wb, file = "ADHD_Screentime_Results.xlsx", overwrite = TRUE)

#CBCL structure
Table_FL_CBCL <- results_CBCL_model[["parameters"]][["stdyx.standardized"]] 
Table_FL_CBCL <- Table_FL_CBCL %>% dplyr::select(paramHeader, param, est, se, pval)
Table_FL_CBCL <- Table_FL_CBCL %>% dplyr::rename(Factor = paramHeader) %>% dplyr::rename(Item = param) %>% dplyr::rename(Loading = est) %>% dplyr::rename(SE = se) %>% dplyr::rename("P-value" = pval)
Table_FL_CBCL <- Table_FL_CBCL[Table_FL_CBCL$Factor != "Thresholds", ]
Table_FL_CBCL <- Table_FL_CBCL[Table_FL_CBCL$Factor != "Variances", ]

wb <- loadWorkbook("ADHD_Screentime_Results.xlsx")
addWorksheet(wb, sheetName = "CBCL 8F structure")
writeData(wb, sheet = "CBCL 8F structure", x = Table_FL_CBCL)
saveWorkbook(wb, file = "ADHD_Screentime_Results.xlsx", overwrite = TRUE)

#SDQ configural
Table_FL_SDQ_invariance_config <- results_SDQ_invariance_testing[["parameters"]][["unstandardized"]][["CONFIGURAL.MODEL"]]
Table_FL_SDQ_invariance_config <- Table_FL_SDQ_invariance_config %>% dplyr::select(paramHeader, param, est, se, pval)
Table_FL_SDQ_invariance_config <- Table_FL_SDQ_invariance_config %>% dplyr::rename(Factor = paramHeader) %>% dplyr::rename(Item = param) %>% dplyr::rename(Loading = est) %>% dplyr::rename(SE = se) %>% dplyr::rename("P-value" = pval)
Table_FL_SDQ_invariance_config <- Table_FL_SDQ_invariance_config[Table_FL_SDQ_invariance_config$Factor != "Thresholds", ]
Table_FL_SDQ_invariance_config <- Table_FL_SDQ_invariance_config[Table_FL_SDQ_invariance_config$Factor != "Variances", ]
Table_FL_SDQ_invariance_config <- Table_FL_SDQ_invariance_config[Table_FL_SDQ_invariance_config$Factor != "Scales", ]
Table_FL_SDQ_invariance_config <- Table_FL_SDQ_invariance_config[Table_FL_SDQ_invariance_config$Factor != "Means", ]

wb <- loadWorkbook("ADHD_Screentime_Results.xlsx")
addWorksheet(wb, sheetName = "SDQ 4F configural model")
writeData(wb, sheet = "SDQ 4F configural model", x = Table_FL_SDQ_invariance_config)
saveWorkbook(wb, file = "ADHD_Screentime_Results.xlsx", overwrite = TRUE)

#SDQ scalar
Table_FL_SDQ_invariance_scalar <- results_SDQ_invariance_testing[["parameters"]][["unstandardized"]][["SCALAR.MODEL"]]
Table_FL_SDQ_invariance_scalar <- Table_FL_SDQ_invariance_scalar %>% dplyr::select(paramHeader, param, est, se, pval)
Table_FL_SDQ_invariance_scalar <- Table_FL_SDQ_invariance_scalar %>% dplyr::rename(Factor = paramHeader) %>% dplyr::rename(Item = param) %>% dplyr::rename(Loading = est) %>% dplyr::rename(SE = se) %>% dplyr::rename("P-value" = pval)
Table_FL_SDQ_invariance_scalar <- Table_FL_SDQ_invariance_scalar[Table_FL_SDQ_invariance_scalar$Factor != "Thresholds", ]
Table_FL_SDQ_invariance_scalar <- Table_FL_SDQ_invariance_scalar[Table_FL_SDQ_invariance_scalar$Factor != "Variances", ]
Table_FL_SDQ_invariance_scalar <- Table_FL_SDQ_invariance_scalar[Table_FL_SDQ_invariance_scalar$Factor != "Scales", ]
Table_FL_SDQ_invariance_scalar <- Table_FL_SDQ_invariance_scalar[Table_FL_SDQ_invariance_scalar$Factor != "Means", ]

wb <- loadWorkbook("ADHD_Screentime_Results.xlsx")
addWorksheet(wb, sheetName = "SDQ 4F scalar model")
writeData(wb, sheet = "SDQ 4F scalar model", x = Table_FL_SDQ_invariance_scalar)
saveWorkbook(wb, file = "ADHD_Screentime_Results.xlsx", overwrite = TRUE)

#CBCL configural
Table_FL_CBCL_ABCL_invariance_config <- results_CBCL_ABCL_invariance_testing[["parameters"]][["unstandardized"]][["CONFIGURAL.MODEL"]]
Table_FL_CBCL_ABCL_invariance_config <- Table_FL_CBCL_ABCL_invariance_config %>% dplyr::select(paramHeader, param, est, se, pval)
Table_FL_CBCL_ABCL_invariance_config <- Table_FL_CBCL_ABCL_invariance_config %>% dplyr::rename(Factor = paramHeader) %>% dplyr::rename(Item = param) %>% dplyr::rename(Loading = est) %>% dplyr::rename(SE = se) %>% dplyr::rename("P-value" = pval)
Table_FL_CBCL_ABCL_invariance_config <- Table_FL_CBCL_ABCL_invariance_config[Table_FL_CBCL_ABCL_invariance_config$Factor != "Thresholds", ]
Table_FL_CBCL_ABCL_invariance_config <- Table_FL_CBCL_ABCL_invariance_config[Table_FL_CBCL_ABCL_invariance_config$Factor != "Variances", ]
Table_FL_CBCL_ABCL_invariance_config <- Table_FL_CBCL_ABCL_invariance_config[Table_FL_CBCL_ABCL_invariance_config$Factor != "Scales", ]
Table_FL_CBCL_ABCL_invariance_config <- Table_FL_CBCL_ABCL_invariance_config[Table_FL_CBCL_ABCL_invariance_config$Factor != "Means", ]

wb <- loadWorkbook("ADHD_Screentime_Results.xlsx")
addWorksheet(wb, sheetName = "CBCL_ABCL configural model")
writeData(wb, sheet = "CBCL_ABCL configural model", x = Table_FL_CBCL_ABCL_invariance_config)
saveWorkbook(wb, file = "ADHD_Screentime_Results.xlsx", overwrite = TRUE)

#CBCL scalar
Table_FL_CBCL_ABCL_invariance_scalar <- results_CBCL_ABCL_invariance_testing[["parameters"]][["unstandardized"]][["SCALAR.MODEL"]]
Table_FL_CBCL_ABCL_invariance_scalar <- Table_FL_CBCL_ABCL_invariance_scalar %>% dplyr::select(paramHeader, param, est, se, pval)
Table_FL_CBCL_ABCL_invariance_scalar <- Table_FL_CBCL_ABCL_invariance_scalar %>% dplyr::rename(Factor = paramHeader) %>% dplyr::rename(Item = param) %>% dplyr::rename(Loading = est) %>% dplyr::rename(SE = se) %>% dplyr::rename("P-value" = pval)
Table_FL_CBCL_ABCL_invariance_scalar <- Table_FL_CBCL_ABCL_invariance_scalar[Table_FL_CBCL_ABCL_invariance_scalar$Factor != "Thresholds", ]
Table_FL_CBCL_ABCL_invariance_scalar <- Table_FL_CBCL_ABCL_invariance_scalar[Table_FL_CBCL_ABCL_invariance_scalar$Factor != "Variances", ]
Table_FL_CBCL_ABCL_invariance_scalar <- Table_FL_CBCL_ABCL_invariance_scalar[Table_FL_CBCL_ABCL_invariance_scalar$Factor != "Scales", ]
Table_FL_CBCL_ABCL_invariance_scalar <- Table_FL_CBCL_ABCL_invariance_scalar[Table_FL_CBCL_ABCL_invariance_scalar$Factor != "Means", ]

wb <- loadWorkbook("ADHD_Screentime_Results.xlsx")
addWorksheet(wb, sheetName = "CBCL_ABCL scalar model")
writeData(wb, sheet = "CBCL_ABCL scalar model", x = Table_FL_CBCL_ABCL_invariance_scalar)
saveWorkbook(wb, file = "ADHD_Screentime_Results.xlsx", overwrite = TRUE)
```

### Descriptive table, Normality and non-parametric/dependent rank tests

```
#Table1 functions
my.render.cont <- function(x) {
    with(stats.apply.rounding(stats.default(x), digits=2), c("",
        "Mean (SD)"=sprintf("%s (&plusmn; %s)", MEAN, SD)))
}


my.render.cat <- function(x) {
    c("", sapply(stats.default(x), function(y) with(y,
        sprintf("%d (%0.1f%%)", FREQ, PCT))))  
}

#Descriptive Table
table_output <- table1(~ Age + Gender + abepstrat + minstlevel_4cat + Maternal_DIAG_ANY + screentime + att_sdq + att_cbcl | wave,
       data = mydata_long,
       render.continuous = my.render.cont,
       render.categorical = my.render.cat)

table_df <- as.data.frame(table_output)

write.xlsx(table_df, file = "Sample_demographics.xlsx", sheetName = "Sample demographics", rowNames = FALSE)

#Normality tests
shapiro.test(mydata_wide$att_w0)
```

```
## 
##  Shapiro-Wilk normality test
## 
## data:  mydata_wide$att_w0
## W = 0.94606, p-value < 2.2e-16
```

```
shapiro.test(mydata_wide$att_w1)
```

```
## 
##  Shapiro-Wilk normality test
## 
## data:  mydata_wide$att_w1
## W = 0.93204, p-value < 2.2e-16
```

```
shapiro.test(mydata_wide$att_w2)
```

```
## 
##  Shapiro-Wilk normality test
## 
## data:  mydata_wide$att_w2
## W = 0.91192, p-value < 2.2e-16
```

```
shapiro.test(mydata_wide$att_cbcl_w0)
```

```
## 
##  Shapiro-Wilk normality test
## 
## data:  mydata_wide$att_cbcl_w0
## W = 0.87739, p-value < 2.2e-16
```

```
shapiro.test(mydata_wide$att_cbcl_w1)
```

```
## 
##  Shapiro-Wilk normality test
## 
## data:  mydata_wide$att_cbcl_w1
## W = 0.8758, p-value < 2.2e-16
```

```
shapiro.test(mydata_wide$att_cbcl_w2)
```

```
## 
##  Shapiro-Wilk normality test
## 
## data:  mydata_wide$att_cbcl_w2
## W = 0.83131, p-value < 2.2e-16
```

```
#Homogeniety tests
wilcox.test(mydata_wide$screentime_w0, mydata_wide$screentime_w1, paired = TRUE)
```

```
## 
##  Wilcoxon signed rank test with continuity correction
## 
## data:  mydata_wide$screentime_w0 and mydata_wide$screentime_w1
## V = 399547, p-value = 6.31e-12
## alternative hypothesis: true location shift is not equal to 0
```

```
wilcox.test(mydata_wide$screentime_w1, mydata_wide$screentime_w2, paired = TRUE)
```

```
## 
##  Wilcoxon signed rank test with continuity correction
## 
## data:  mydata_wide$screentime_w1 and mydata_wide$screentime_w2
## V = 219332, p-value = 0.0003273
## alternative hypothesis: true location shift is not equal to 0
```

```
wilcox.test(mydata_wide$att_w0, mydata_wide$att_w1, paired = TRUE)
```

```
## 
##  Wilcoxon signed rank test with continuity correction
## 
## data:  mydata_wide$att_w0 and mydata_wide$att_w1
## V = 936216, p-value < 2.2e-16
## alternative hypothesis: true location shift is not equal to 0
```

```
wilcox.test(mydata_wide$att_w1, mydata_wide$att_w2, paired = TRUE)
```

```
## 
##  Wilcoxon signed rank test with continuity correction
## 
## data:  mydata_wide$att_w1 and mydata_wide$att_w2
## V = 499011, p-value = 1.398e-15
## alternative hypothesis: true location shift is not equal to 0
```

```
wilcox.test(mydata_wide$att_cbcl_w0, mydata_wide$att_cbcl_w1, paired = TRUE)
```

```
## 
##  Wilcoxon signed rank test with continuity correction
## 
## data:  mydata_wide$att_cbcl_w0 and mydata_wide$att_cbcl_w1
## V = 703345, p-value = 0.01098
## alternative hypothesis: true location shift is not equal to 0
```

```
wilcox.test(mydata_wide$att_cbcl_w1, mydata_wide$att_cbcl_w2, paired = TRUE)
```

```
## 
##  Wilcoxon signed rank test with continuity correction
## 
## data:  mydata_wide$att_cbcl_w1 and mydata_wide$att_cbcl_w2
## V = 363501, p-value = 8.999e-13
## alternative hypothesis: true location shift is not equal to 0
```

### SDQ Random Intercept Cross-Lagged Models

```
# Random intercept CLPM
ri_clpm_screentime_AttSDQ <- '
# Random intercepts
RI_att =~ 1*att_w0 + 1*att_w1 + 1*att_w2
RI_scr =~ 1*screentime_ord_w0 + 1*screentime_ord_w1 + 1*screentime_ord_w2

# Estimate variance and covariance of random intercepts
RI_att ~~ RI_att
RI_scr ~~ RI_scr
RI_att ~~ RI_scr

# Observed var intercepts (triangles on schematic plots)
att_w0 ~ iat1*1 
att_w1 ~ iat2*1
att_w2 ~ iat3*1
screentime_ord_w0 ~ ist1*1
screentime_ord_w1 ~ ist2*1
screentime_ord_w2 ~ ist3*1

# Create within-person centered latent variables
l_att_w0 =~ 1*att_w0
l_att_w1 =~ 1*att_w1
l_att_w2 =~ 1*att_w2
l_scr_w0 =~ 1*screentime_ord_w0 
l_scr_w1 =~ 1*screentime_ord_w1 
l_scr_w2 =~ 1*screentime_ord_w2 

# Estimate lagged effects between within-person centered variables
## Autoregressive paths
l_scr_w1 ~ l_scr_w0
l_scr_w2 ~ l_scr_w1
l_att_w1 ~ l_att_w0 
l_att_w2 ~ l_att_w1
## Cross-lagged paths
l_scr_w1 ~ l_att_w0 
l_scr_w2 ~ l_att_w1 
l_att_w1 ~ l_scr_w0
l_att_w2 ~ l_scr_w1

# Estimate (residual) variance of within-person centered exogenous variables
l_att_w0 ~~ l_att_w0 
l_att_w1 ~~ l_att_w1
l_att_w2 ~~ l_att_w2 
l_scr_w0 ~~ l_scr_w0 
l_scr_w1 ~~ l_scr_w1
l_scr_w2 ~~ l_scr_w2 

# Estimate covariance between within-person centered exogenous variables
l_att_w0 ~~ l_scr_w0
l_att_w1 ~~ l_scr_w1
l_att_w2 ~~ l_scr_w2 
'


sem_ri_clpm_screen_AttSDQ <- lavaan(ri_clpm_screentime_AttSDQ, data=mydata_wide, std.lv = F,
                                 missing="ML", sampling.weights = "sampling_ipw", 
                                 int.ov.free = F, int.lv.free = F, auto.fix.first = F, auto.fix.single = F, 
                                 auto.cov.lv.x = F, auto.cov.y = F, auto.var = F)

summary(sem_ri_clpm_screen_AttSDQ, fit.measures=T, standardized=T, rsquare=T)
```

```
## lavaan 0.6-19 ended normally after 207 iterations
## 
##   Estimator                                         ML
##   Optimization method                           NLMINB
##   Number of model parameters                        26
## 
##   Number of observations                              2511
##   Number of missing patterns                            18
##   Sampling weights variable                   sampling_ipw
## 
## Model Test User Model:
##                                               Standard      Scaled
##   Test Statistic                                 1.664       0.921
##   Degrees of freedom                                 1           1
##   P-value (Chi-square)                           0.197       0.337
##   Scaling correction factor                                  1.808
##     Yuan-Bentler correction (Mplus variant)                       
## 
## Model Test Baseline Model:
## 
##   Test statistic                              1350.395     758.825
##   Degrees of freedom                                15          15
##   P-value                                        0.000       0.000
##   Scaling correction factor                                  1.780
## 
## User Model versus Baseline Model:
## 
##   Comparative Fit Index (CFI)                    1.000       1.000
##   Tucker-Lewis Index (TLI)                       0.993       1.002
##                                                                   
##   Robust Comparative Fit Index (CFI)                         1.000
##   Robust Tucker-Lewis Index (TLI)                            1.002
## 
## Loglikelihood and Information Criteria:
## 
##   Loglikelihood user model (H0)             -25871.474  -25871.474
##   Scaling correction factor                                  1.580
##       for the MLR correction                                      
##   Loglikelihood unrestricted model (H1)     -25870.642  -25870.642
##   Scaling correction factor                                  1.589
##       for the MLR correction                                      
##                                                                   
##   Akaike (AIC)                               51794.948   51794.948
##   Bayesian (BIC)                             51946.488   51946.488
##   Sample-size adjusted Bayesian (SABIC)      51863.879   51863.879
## 
## Root Mean Square Error of Approximation:
## 
##   RMSEA                                          0.016       0.000
##   90 Percent confidence interval - lower         0.000       0.000
##   90 Percent confidence interval - upper         0.059       0.040
##   P-value H_0: RMSEA <= 0.050                    0.888       0.989
##   P-value H_0: RMSEA >= 0.080                    0.003       0.000
##                                                                   
##   Robust RMSEA                                               0.000
##   90 Percent confidence interval - lower                     0.000
##   90 Percent confidence interval - upper                     0.099
##   P-value H_0: Robust RMSEA <= 0.050                         0.649
##   P-value H_0: Robust RMSEA >= 0.080                         0.125
## 
## Standardized Root Mean Square Residual:
## 
##   SRMR                                           0.007       0.007
## 
## Parameter Estimates:
## 
##   Standard errors                             Sandwich
##   Information bread                           Observed
##   Observed information based on                Hessian
## 
## Latent Variables:
##                    Estimate  Std.Err  z-value  P(>|z|)   Std.lv  Std.all
##   RI_att =~                                                             
##     att_w0            1.000                               1.830    0.607
##     att_w1            1.000                               1.830    0.611
##     att_w2            1.000                               1.830    0.640
##   RI_scr =~                                                             
##     screentm_rd_w0    1.000                               0.776    0.448
##     screentm_rd_w1    1.000                               0.776    0.384
##     screentm_rd_w2    1.000                               0.776    0.360
##   l_att_w0 =~                                                           
##     att_w0            1.000                               2.393    0.794
##   l_att_w1 =~                                                           
##     att_w1            1.000                               2.371    0.792
##   l_att_w2 =~                                                           
##     att_w2            1.000                               2.199    0.769
##   l_scr_w0 =~                                                           
##     screentm_rd_w0    1.000                               1.548    0.894
##   l_scr_w1 =~                                                           
##     screentm_rd_w1    1.000                               1.864    0.923
##   l_scr_w2 =~                                                           
##     screentm_rd_w2    1.000                               2.009    0.933
## 
## Regressions:
##                    Estimate  Std.Err  z-value  P(>|z|)   Std.lv  Std.all
##   l_scr_w1 ~                                                            
##     l_scr_w0         -0.016    0.075   -0.218    0.828   -0.014   -0.014
##   l_scr_w2 ~                                                            
##     l_scr_w1         -0.049    0.068   -0.721    0.471   -0.045   -0.045
##   l_att_w1 ~                                                            
##     l_att_w0          0.215    0.056    3.807    0.000    0.217    0.217
##   l_att_w2 ~                                                            
##     l_att_w1          0.214    0.060    3.580    0.000    0.231    0.231
##   l_scr_w1 ~                                                            
##     l_att_w0         -0.046    0.040   -1.145    0.252   -0.059   -0.059
##   l_scr_w2 ~                                                            
##     l_att_w1         -0.002    0.047   -0.036    0.972   -0.002   -0.002
##   l_att_w1 ~                                                            
##     l_scr_w0          0.019    0.070    0.278    0.781    0.013    0.013
##   l_att_w2 ~                                                            
##     l_scr_w1          0.010    0.054    0.177    0.859    0.008    0.008
## 
## Covariances:
##                    Estimate  Std.Err  z-value  P(>|z|)   Std.lv  Std.all
##   RI_att ~~                                                             
##     RI_scr            0.337    0.150    2.238    0.025    0.237    0.237
##   l_att_w0 ~~                                                           
##     l_scr_w0          0.248    0.178    1.398    0.162    0.067    0.067
##  .l_att_w1 ~~                                                           
##    .l_scr_w1          0.352    0.194    1.816    0.069    0.082    0.082
##  .l_att_w2 ~~                                                           
##    .l_scr_w2          0.296    0.205    1.444    0.149    0.069    0.069
## 
## Intercepts:
##                    Estimate  Std.Err  z-value  P(>|z|)   Std.lv  Std.all
##    .att_w0  (iat1)    4.436    0.078   56.977    0.000    4.436    1.473
##    .att_w1  (iat2)    3.623    0.084   42.959    0.000    3.623    1.210
##    .att_w2  (iat3)    3.164    0.085   37.205    0.000    3.164    1.106
##    .scrn__0 (ist1)    3.253    0.047   68.687    0.000    3.253    1.878
##    .scrn__1 (ist2)    3.543    0.062   57.432    0.000    3.543    1.754
##    .scrn__2 (ist3)    3.198    0.082   39.077    0.000    3.198    1.484
## 
## Variances:
##                    Estimate  Std.Err  z-value  P(>|z|)   Std.lv  Std.all
##     RI_att            3.348    0.341    9.828    0.000    1.000    1.000
##     RI_scr            0.603    0.143    4.214    0.000    1.000    1.000
##     l_att_w0          5.724    0.363   15.757    0.000    1.000    1.000
##    .l_att_w1          5.354    0.322   16.619    0.000    0.953    0.953
##    .l_att_w2          4.575    0.277   16.497    0.000    0.946    0.946
##     l_scr_w0          2.397    0.159   15.047    0.000    1.000    1.000
##    .l_scr_w1          3.463    0.200   17.351    0.000    0.996    0.996
##    .l_scr_w2          4.029    0.217   18.533    0.000    0.998    0.998
##    .att_w0            0.000                               0.000    0.000
##    .att_w1            0.000                               0.000    0.000
##    .att_w2            0.000                               0.000    0.000
##    .screentm_rd_w0    0.000                               0.000    0.000
##    .screentm_rd_w1    0.000                               0.000    0.000
##    .screentm_rd_w2    0.000                               0.000    0.000
## 
## R-Square:
##                    Estimate
##     l_att_w1          0.047
##     l_att_w2          0.054
##     l_scr_w1          0.004
##     l_scr_w2          0.002
##     att_w0            1.000
##     att_w1            1.000
##     att_w2            1.000
##     screentm_rd_w0    1.000
##     screentm_rd_w1    1.000
##     screentm_rd_w2    1.000
```

```
parameterEstimates(sem_ri_clpm_screen_AttSDQ, ci = TRUE, level = 0.95, boot.ci.type = "perc", standardized = TRUE) %>% 
  filter(op == "~") %>% 
  dplyr::select('Regressions'=lhs, Indicator=rhs, B=est, SE=se, Z=z, 'p-value'=pvalue, Beta=std.all) %>% 
  kable(digits = 3, format="pandoc", caption="Regression coeficients from CLPM")
```

Regression coeficients from CLPM

| Regressions | Indicator | B | SE | Z | p-value | Beta |
| --- | --- | --- | --- | --- | --- | --- |
| l\_scr\_w1 | l\_scr\_w0 | -0.016 | 0.075 | -0.218 | 0.828 | -0.014 |
| l\_scr\_w2 | l\_scr\_w1 | -0.049 | 0.068 | -0.721 | 0.471 | -0.045 |
| l\_att\_w1 | l\_att\_w0 | 0.215 | 0.056 | 3.807 | 0.000 | 0.217 |
| l\_att\_w2 | l\_att\_w1 | 0.214 | 0.060 | 3.580 | 0.000 | 0.231 |
| l\_scr\_w1 | l\_att\_w0 | -0.046 | 0.040 | -1.145 | 0.252 | -0.059 |
| l\_scr\_w2 | l\_att\_w1 | -0.002 | 0.047 | -0.036 | 0.972 | -0.002 |
| l\_att\_w1 | l\_scr\_w0 | 0.019 | 0.070 | 0.278 | 0.781 | 0.013 |
| l\_att\_w2 | l\_scr\_w1 | 0.010 | 0.054 | 0.177 | 0.859 | 0.008 |

```
#Standardized
Paths_sem_ri_clpm_screen_AttSDQ <- standardizedsolution(sem_ri_clpm_screen_AttSDQ, ci = TRUE, level = 0.95) %>% 
  filter(op != "=~") %>% 
  mutate(
    Beta_CI = sprintf("%.3f (%.3f to %.3f)", est.std, ci.lower, ci.upper),
    SE = sprintf("%.3f", se),
    Z = sprintf("%.3f", z),
    pvalue = sprintf("%.3f", pvalue),
    Beta = sprintf("%.3f", est.std)
  ) %>%
  dplyr::select('Regressions' = lhs, Indicator = rhs, Beta_CI, SE, Z, 'p-value' = pvalue, Beta)

Paths_sem_ri_clpm_screen_AttSDQ <- Paths_sem_ri_clpm_screen_AttSDQ[3:26,]
Paths_sem_ri_clpm_screen_AttSDQ <- Paths_sem_ri_clpm_screen_AttSDQ[-2:-7,]
Paths_sem_ri_clpm_screen_AttSDQ <- Paths_sem_ri_clpm_screen_AttSDQ[-10:-15,]

wb <- loadWorkbook("ADHD_Screentime_Results.xlsx")
addWorksheet(wb, sheetName = "RI-CLPM Screen-AttSDQ")
writeData(wb, sheet = "RI-CLPM Screen-AttSDQ", x = Paths_sem_ri_clpm_screen_AttSDQ)
saveWorkbook(wb, file = "ADHD_Screentime_Results.xlsx", overwrite = TRUE)

# Random intercept CLPM, adjusted
ri_clpm_screentime_AttSDQ_adjust <- '
# Random intercepts
RI_att =~ 1*att_w0 + 1*att_w1 + 1*att_w2
RI_scr =~ 1*screentime_ord_w0 + 1*screentime_ord_w1 + 1*screentime_ord_w2

# Estimate variance and covariance of random intercepts
RI_att ~~ RI_att
RI_scr ~~ RI_scr
RI_att ~~ RI_scr

# Observed var intercepts (triangles on schematic plots)
att_w0 ~ iat1*1 
att_w1 ~ iat2*1
att_w2 ~ iat3*1
screentime_ord_w0 ~ ist1*1
screentime_ord_w1 ~ ist2*1
screentime_ord_w2 ~ ist3*1

# Create within-person centered latent variables
l_att_w0 =~ 1*att_w0
l_att_w1 =~ 1*att_w1
l_att_w2 =~ 1*att_w2
l_scr_w0 =~ 1*screentime_ord_w0 
l_scr_w1 =~ 1*screentime_ord_w1 
l_scr_w2 =~ 1*screentime_ord_w2 

# Estimate lagged effects between within-person centered variables
## Autoregressive paths
l_scr_w1 ~ l_scr_w0
l_scr_w2 ~ l_scr_w1
l_att_w1 ~ l_att_w0 
l_att_w2 ~ l_att_w1
## Cross-lagged paths
l_scr_w1 ~ l_att_w0 
l_scr_w2 ~ l_att_w1 
l_att_w1 ~ l_scr_w0
l_att_w2 ~ l_scr_w1

# Estimate (residual) variance of within-person centered exogenous variables
l_att_w0 ~~ l_att_w0 
l_att_w1 ~~ l_att_w1
l_att_w2 ~~ l_att_w2 
l_scr_w0 ~~ l_scr_w0 
l_scr_w1 ~~ l_scr_w1
l_scr_w2 ~~ l_scr_w2 

# Estimate covariance between within-person centered exogenous variables
l_att_w0 ~~ l_scr_w0
l_att_w1 ~~ l_scr_w1
l_att_w2 ~~ l_scr_w2 

# Adjusted
RI_att ~ Age_w0_z + Gender + abepstrat_w0 + minstlevel_4cat_w0 + Maternal_DIAG_ANY
RI_scr ~ Age_w0_z + Gender + abepstrat_w0 + minstlevel_4cat_w0 + Maternal_DIAG_ANY

# Variance and Covariance of covariates
Age_w0_z ~~ Age_w0_z
Gender ~~ Gender
abepstrat_w0 ~~ abepstrat_w0
minstlevel_4cat_w0 ~~ minstlevel_4cat_w0
Maternal_DIAG_ANY ~~ Maternal_DIAG_ANY

Age_w0_z ~~ Gender
Age_w0_z ~~ abepstrat_w0
Age_w0_z ~~ Maternal_DIAG_ANY
Age_w0_z ~~ minstlevel_4cat_w0
Gender ~~ abepstrat_w0
Gender ~~ Maternal_DIAG_ANY
Gender ~~ minstlevel_4cat_w0
abepstrat_w0 ~~ Maternal_DIAG_ANY
abepstrat_w0 ~~ minstlevel_4cat_w0
minstlevel_4cat_w0 ~~ Maternal_DIAG_ANY


'


sem_ri_clpm_screen_AttSDQ_adjust <- lavaan(ri_clpm_screentime_AttSDQ_adjust, data=mydata_wide, std.lv = F, 
                                 missing="ML", sampling.weights = "sampling_ipw", 
                                 int.ov.free = F, int.lv.free = F, auto.fix.first = F, auto.fix.single = F, 
                                 auto.cov.lv.x = F, auto.cov.y = F, auto.var = F,
                                 std.ov=T) 

summary(sem_ri_clpm_screen_AttSDQ_adjust, fit.measures=T, standardized=T, rsquare=T)
```

```
## lavaan 0.6-19 ended normally after 42 iterations
## 
##   Estimator                                         ML
##   Optimization method                           NLMINB
##   Number of model parameters                        51
## 
##   Number of observations                              2511
##   Number of missing patterns                            30
##   Sampling weights variable                   sampling_ipw
## 
## Model Test User Model:
##                                               Standard      Scaled
##   Test Statistic                               179.804     105.124
##   Degrees of freedom                                26          26
##   P-value (Chi-square)                           0.000       0.000
##   Scaling correction factor                                  1.710
##     Yuan-Bentler correction (Mplus variant)                       
## 
## Model Test Baseline Model:
## 
##   Test statistic                              1854.853    1086.024
##   Degrees of freedom                                55          55
##   P-value                                        0.000       0.000
##   Scaling correction factor                                  1.708
## 
## User Model versus Baseline Model:
## 
##   Comparative Fit Index (CFI)                    0.915       0.923
##   Tucker-Lewis Index (TLI)                       0.819       0.838
##                                                                   
##   Robust Comparative Fit Index (CFI)                         0.931
##   Robust Tucker-Lewis Index (TLI)                            0.855
## 
## Loglikelihood and Information Criteria:
## 
##   Loglikelihood user model (H0)             -32698.091  -32698.091
##   Scaling correction factor                                  1.557
##       for the MLR correction                                      
##   Loglikelihood unrestricted model (H1)     -32608.190  -32608.190
##   Scaling correction factor                                  1.609
##       for the MLR correction                                      
##                                                                   
##   Akaike (AIC)                               65498.183   65498.183
##   Bayesian (BIC)                             65795.433   65795.433
##   Sample-size adjusted Bayesian (SABIC)      65633.393   65633.393
## 
## Root Mean Square Error of Approximation:
## 
##   RMSEA                                          0.049       0.035
##   90 Percent confidence interval - lower         0.042       0.030
##   90 Percent confidence interval - upper         0.055       0.040
##   P-value H_0: RMSEA <= 0.050                    0.626       1.000
##   P-value H_0: RMSEA >= 0.080                    0.000       0.000
##                                                                   
##   Robust RMSEA                                               0.050
##   90 Percent confidence interval - lower                     0.039
##   90 Percent confidence interval - upper                     0.061
##   P-value H_0: Robust RMSEA <= 0.050                         0.494
##   P-value H_0: Robust RMSEA >= 0.080                         0.000
## 
## Standardized Root Mean Square Residual:
## 
##   SRMR                                           0.033       0.033
## 
## Parameter Estimates:
## 
##   Standard errors                             Sandwich
##   Information bread                           Observed
##   Observed information based on                Hessian
## 
## Latent Variables:
##                    Estimate  Std.Err  z-value  P(>|z|)   Std.lv  Std.all
##   RI_att =~                                                             
##     att_w0            1.000                               0.608    0.627
##     att_w1            1.000                               0.608    0.627
##     att_w2            1.000                               0.608    0.629
##   RI_scr =~                                                             
##     screentm_rd_w0    1.000                               0.402    0.411
##     screentm_rd_w1    1.000                               0.402    0.411
##     screentm_rd_w2    1.000                               0.402    0.403
##   l_att_w0 =~                                                           
##     att_w0            1.000                               0.756    0.779
##   l_att_w1 =~                                                           
##     att_w1            1.000                               0.756    0.779
##   l_att_w2 =~                                                           
##     att_w2            1.000                               0.751    0.777
##   l_scr_w0 =~                                                           
##     screentm_rd_w0    1.000                               0.890    0.912
##   l_scr_w1 =~                                                           
##     screentm_rd_w1    1.000                               0.891    0.912
##   l_scr_w2 =~                                                           
##     screentm_rd_w2    1.000                               0.911    0.915
## 
## Regressions:
##                    Estimate  Std.Err  z-value  P(>|z|)   Std.lv  Std.all
##   l_scr_w1 ~                                                            
##     l_scr_w0         -0.014    0.058   -0.235    0.815   -0.014   -0.014
##   l_scr_w2 ~                                                            
##     l_scr_w1         -0.073    0.070   -1.036    0.300   -0.071   -0.071
##   l_att_w1 ~                                                            
##     l_att_w0          0.183    0.060    3.049    0.002    0.183    0.183
##   l_att_w2 ~                                                            
##     l_att_w1          0.231    0.059    3.907    0.000    0.233    0.233
##   l_scr_w1 ~                                                            
##     l_att_w0         -0.068    0.063   -1.090    0.276   -0.058   -0.058
##   l_scr_w2 ~                                                            
##     l_att_w1         -0.030    0.069   -0.439    0.661   -0.025   -0.025
##   l_att_w1 ~                                                            
##     l_scr_w0          0.016    0.038    0.432    0.666    0.019    0.019
##   l_att_w2 ~                                                            
##     l_scr_w1          0.016    0.038    0.412    0.680    0.019    0.019
##   RI_att ~                                                              
##     Age_w0_z         -0.094    0.021   -4.513    0.000   -0.154   -0.155
##     Gender            0.112    0.020    5.498    0.000    0.184    0.185
##     abepstrat_w0     -0.034    0.022   -1.534    0.125   -0.055   -0.054
##     mnstlvl_4ct_w0    0.008    0.022    0.348    0.727    0.013    0.013
##     Mtrnl_DIAG_ANY    0.178    0.021    8.447    0.000    0.292    0.272
##   RI_scr ~                                                              
##     Age_w0_z          0.035    0.020    1.790    0.073    0.087    0.087
##     Gender            0.027    0.019    1.406    0.160    0.068    0.068
##     abepstrat_w0      0.040    0.019    2.080    0.038    0.099    0.097
##     mnstlvl_4ct_w0    0.053    0.020    2.641    0.008    0.131    0.131
##     Mtrnl_DIAG_ANY    0.067    0.019    3.539    0.000    0.168    0.156
## 
## Covariances:
##                         Estimate  Std.Err  z-value  P(>|z|)   Std.lv  Std.all
##  .RI_att ~~                                                                  
##    .RI_scr                 0.050    0.025    1.983    0.047    0.225    0.225
##   l_att_w0 ~~                                                                
##     l_scr_w0               0.047    0.031    1.484    0.138    0.069    0.069
##  .l_att_w1 ~~                                                                
##    .l_scr_w1               0.057    0.032    1.778    0.075    0.086    0.086
##  .l_att_w2 ~~                                                                
##    .l_scr_w2               0.039    0.032    1.220    0.222    0.059    0.059
##   Age_w0_z ~~                                                                
##     Gender                -0.035    0.027   -1.275    0.202   -0.035   -0.034
##     abepstrat_w0           0.030    0.026    1.126    0.260    0.030    0.030
##     Mtrnl_DIAG_ANY         0.043    0.024    1.769    0.077    0.043    0.046
##     mnstlvl_4ct_w0        -0.072    0.026   -2.733    0.006   -0.072   -0.072
##   Gender ~~                                                                  
##     abepstrat_w0           0.043    0.025    1.713    0.087    0.043    0.043
##     Mtrnl_DIAG_ANY        -0.036    0.024   -1.498    0.134   -0.036   -0.038
##     mnstlvl_4ct_w0         0.090    0.026    3.419    0.001    0.090    0.090
##   abepstrat_w0 ~~                                                            
##     Mtrnl_DIAG_ANY        -0.037    0.023   -1.614    0.107   -0.037   -0.040
##     mnstlvl_4ct_w0         0.233    0.025    9.344    0.000    0.233    0.238
##   minstlevel_4cat_w0 ~~                                                      
##     Mtrnl_DIAG_ANY        -0.005    0.024   -0.214    0.831   -0.005   -0.005
## 
## Intercepts:
##                    Estimate  Std.Err  z-value  P(>|z|)   Std.lv  Std.all
##    .att_w0  (iat1)   -0.129    0.024   -5.287    0.000   -0.129   -0.133
##    .att_w1  (iat2)   -0.132    0.027   -4.920    0.000   -0.132   -0.136
##    .att_w2  (iat3)   -0.094    0.028   -3.315    0.001   -0.094   -0.097
##    .scrn__0 (ist1)   -0.011    0.026   -0.419    0.675   -0.011   -0.011
##    .scrn__1 (ist2)   -0.024    0.030   -0.800    0.424   -0.024   -0.024
##    .scrn__2 (ist3)   -0.030    0.037   -0.797    0.426   -0.030   -0.030
## 
## Variances:
##                    Estimate  Std.Err  z-value  P(>|z|)   Std.lv  Std.all
##    .RI_att            0.322    0.037    8.745    0.000    0.869    0.869
##    .RI_scr            0.150    0.035    4.244    0.000    0.931    0.931
##     l_att_w0          0.572    0.038   14.894    0.000    1.000    1.000
##    .l_att_w1          0.552    0.034   16.172    0.000    0.966    0.966
##    .l_att_w2          0.534    0.029   18.092    0.000    0.945    0.945
##     l_scr_w0          0.793    0.044   18.149    0.000    1.000    1.000
##    .l_scr_w1          0.790    0.048   16.445    0.000    0.996    0.996
##    .l_scr_w2          0.825    0.051   16.076    0.000    0.994    0.994
##     Age_w0_z          1.005    0.028   36.327    0.000    1.005    1.000
##     Gender            1.010    0.005  199.247    0.000    1.010    1.000
##     abepstrat_w0      0.967    0.040   24.119    0.000    0.967    1.000
##     mnstlvl_4ct_w0    0.995    0.025   39.636    0.000    0.995    1.000
##     Mtrnl_DIAG_ANY    0.865    0.021   41.825    0.000    0.865    1.000
##    .att_w0            0.000                               0.000    0.000
##    .att_w1            0.000                               0.000    0.000
##    .att_w2            0.000                               0.000    0.000
##    .screentm_rd_w0    0.000                               0.000    0.000
##    .screentm_rd_w1    0.000                               0.000    0.000
##    .screentm_rd_w2    0.000                               0.000    0.000
## 
## R-Square:
##                    Estimate
##     RI_att            0.131
##     RI_scr            0.069
##     l_att_w1          0.034
##     l_att_w2          0.055
##     l_scr_w1          0.004
##     l_scr_w2          0.006
##     att_w0            1.000
##     att_w1            1.000
##     att_w2            1.000
##     screentm_rd_w0    1.000
##     screentm_rd_w1    1.000
##     screentm_rd_w2    1.000
```

```
parameterEstimates(sem_ri_clpm_screen_AttSDQ_adjust, ci = TRUE, level = 0.95, boot.ci.type = "perc", standardized = TRUE) %>% 
  filter(op == "~") %>% 
  dplyr::select('Regressions'=lhs, Indicator=rhs, B=est, SE=se, Z=z, 'p-value'=pvalue, Beta=std.all) %>% 
  kable(digits = 3, format="pandoc", caption="Regression coeficients from CLPM")
```

Regression coeficients from CLPM

| Regressions | Indicator | B | SE | Z | p-value | Beta |
| --- | --- | --- | --- | --- | --- | --- |
| l\_scr\_w1 | l\_scr\_w0 | -0.014 | 0.058 | -0.235 | 0.815 | -0.014 |
| l\_scr\_w2 | l\_scr\_w1 | -0.073 | 0.070 | -1.036 | 0.300 | -0.071 |
| l\_att\_w1 | l\_att\_w0 | 0.183 | 0.060 | 3.049 | 0.002 | 0.183 |
| l\_att\_w2 | l\_att\_w1 | 0.231 | 0.059 | 3.907 | 0.000 | 0.233 |
| l\_scr\_w1 | l\_att\_w0 | -0.068 | 0.063 | -1.090 | 0.276 | -0.058 |
| l\_scr\_w2 | l\_att\_w1 | -0.030 | 0.069 | -0.439 | 0.661 | -0.025 |
| l\_att\_w1 | l\_scr\_w0 | 0.016 | 0.038 | 0.432 | 0.666 | 0.019 |
| l\_att\_w2 | l\_scr\_w1 | 0.016 | 0.038 | 0.412 | 0.680 | 0.019 |
| RI\_att | Age\_w0\_z | -0.094 | 0.021 | -4.513 | 0.000 | -0.155 |
| RI\_att | Gender | 0.112 | 0.020 | 5.498 | 0.000 | 0.185 |
| RI\_att | abepstrat\_w0 | -0.034 | 0.022 | -1.534 | 0.125 | -0.054 |
| RI\_att | minstlevel\_4cat\_w0 | 0.008 | 0.022 | 0.348 | 0.727 | 0.013 |
| RI\_att | Maternal\_DIAG\_ANY | 0.178 | 0.021 | 8.447 | 0.000 | 0.272 |
| RI\_scr | Age\_w0\_z | 0.035 | 0.020 | 1.790 | 0.073 | 0.087 |
| RI\_scr | Gender | 0.027 | 0.019 | 1.406 | 0.160 | 0.068 |
| RI\_scr | abepstrat\_w0 | 0.040 | 0.019 | 2.080 | 0.038 | 0.097 |
| RI\_scr | minstlevel\_4cat\_w0 | 0.053 | 0.020 | 2.641 | 0.008 | 0.131 |
| RI\_scr | Maternal\_DIAG\_ANY | 0.067 | 0.019 | 3.539 | 0.000 | 0.156 |

```
#Standardized
Paths_sem_ri_clpm_screen_AttSDQ_adjust <- standardizedsolution(sem_ri_clpm_screen_AttSDQ_adjust, ci = TRUE, level = 0.95) %>% 
  filter(op != "=~") %>% 
  mutate(
    Beta_CI = sprintf("%.3f (%.3f to %.3f)", est.std, ci.lower, ci.upper),
    SE = sprintf("%.3f", se),
    Z = sprintf("%.3f", z),
    pvalue = sprintf("%.3f", pvalue),
    Beta = sprintf("%.3f", est.std)
  ) %>%
  dplyr::select('Regressions' = lhs, Indicator = rhs, Beta_CI, SE, Z, 'p-value' = pvalue, Beta)

Paths_sem_ri_clpm_screen_AttSDQ_adjust <- Paths_sem_ri_clpm_screen_AttSDQ_adjust[3:44,]
Paths_sem_ri_clpm_screen_AttSDQ_adjust <- Paths_sem_ri_clpm_screen_AttSDQ_adjust[-2:-7,]
Paths_sem_ri_clpm_screen_AttSDQ_adjust <- Paths_sem_ri_clpm_screen_AttSDQ_adjust[-10:-15,]
Paths_sem_ri_clpm_screen_AttSDQ_adjust <- Paths_sem_ri_clpm_screen_AttSDQ_adjust[-23:-30,]

wb <- loadWorkbook("ADHD_Screentime_Results.xlsx")
addWorksheet(wb, sheetName = "Adj RI-CLPM Screen-AttSDQ")
writeData(wb, sheet = "Adj RI-CLPM Screen-AttSDQ", x = Paths_sem_ri_clpm_screen_AttSDQ_adjust)
saveWorkbook(wb, file = "ADHD_Screentime_Results.xlsx", overwrite = TRUE)
```

### Sensitivity Analysis

```
# Random intercept CLPM, adjusted
ri_clpm_screentime_AttCBCL_adjust <- '
# Random intercepts
RI_att =~ 1*att_cbcl_w0 + 1*att_cbcl_w1 + 1*att_cbcl_w2
RI_scr =~ 1*screentime_ord_w0 + 1*screentime_ord_w1 + 1*screentime_ord_w2

# Estimate variance and covariance of random intercepts
RI_att ~~ RI_att
RI_scr ~~ RI_scr
RI_att ~~ RI_scr

# Observed var intercepts (triangles on schematic plots)
att_cbcl_w0 ~ iat1*1 
att_cbcl_w1 ~ iat2*1
att_cbcl_w2 ~ iat3*1
screentime_ord_w0 ~ ist1*1
screentime_ord_w1 ~ ist2*1
screentime_ord_w2 ~ ist3*1

# Create within-person centered latent variables
l_att_w0 =~ 1*att_cbcl_w0
l_att_w1 =~ 1*att_cbcl_w1
l_att_w2 =~ 1*att_cbcl_w2
l_scr_w0 =~ 1*screentime_ord_w0 
l_scr_w1 =~ 1*screentime_ord_w1 
l_scr_w2 =~ 1*screentime_ord_w2 

# Estimate lagged effects between within-person centered variables
## Autoregressive paths
l_scr_w1 ~ l_scr_w0
l_scr_w2 ~ l_scr_w1
l_att_w1 ~ l_att_w0 
l_att_w2 ~ l_att_w1
## Cross-lagged paths
l_scr_w1 ~ l_att_w0 
l_scr_w2 ~ l_att_w1 
l_att_w1 ~ l_scr_w0
l_att_w2 ~ l_scr_w1

# Estimate (residual) variance of within-person centered exogenous variables
l_att_w0 ~~ l_att_w0 
l_att_w1 ~~ l_att_w1
l_att_w2 ~~ l_att_w2 
l_scr_w0 ~~ l_scr_w0 
l_scr_w1 ~~ l_scr_w1
l_scr_w2 ~~ l_scr_w2 

# Estimate covariance between within-person centered exogenous variables
l_att_w0 ~~ l_scr_w0
l_att_w1 ~~ l_scr_w1
l_att_w2 ~~ l_scr_w2 

# Adjusted
RI_att ~ Age_w0_z + Gender + abepstrat_w0 + minstlevel_4cat_w0 + Maternal_DIAG_ANY
RI_scr ~ Age_w0_z + Gender + abepstrat_w0 + minstlevel_4cat_w0 + Maternal_DIAG_ANY

# Variance and Covariance of covariates
Age_w0_z ~~ Age_w0_z
Gender ~~ Gender
abepstrat_w0 ~~ abepstrat_w0
minstlevel_4cat_w0 ~~ minstlevel_4cat_w0
Maternal_DIAG_ANY ~~ Maternal_DIAG_ANY

Age_w0_z ~~ Gender
Age_w0_z ~~ abepstrat_w0
Age_w0_z ~~ Maternal_DIAG_ANY
Age_w0_z ~~ minstlevel_4cat_w0
Gender ~~ abepstrat_w0
Gender ~~ Maternal_DIAG_ANY
Gender ~~ minstlevel_4cat_w0
abepstrat_w0 ~~ Maternal_DIAG_ANY
abepstrat_w0 ~~ minstlevel_4cat_w0
minstlevel_4cat_w0 ~~ Maternal_DIAG_ANY
'


sem_ri_clpm_screen_AttCBCL_adjust <- lavaan(ri_clpm_screentime_AttCBCL_adjust, data=mydata_wide, std.lv = F, 
                                 missing="ML", sampling.weights = "sampling_ipw", 
                                 int.ov.free = F, int.lv.free = F, auto.fix.first = F, auto.fix.single = F, 
                                 auto.cov.lv.x = F, auto.cov.y = F, auto.var = F,
                                 std.ov=T) 

summary(sem_ri_clpm_screen_AttCBCL_adjust, fit.measures=T, standardized=T, rsquare=T)
```

```
## lavaan 0.6-19 ended normally after 41 iterations
## 
##   Estimator                                         ML
##   Optimization method                           NLMINB
##   Number of model parameters                        51
## 
##   Number of observations                              2511
##   Number of missing patterns                            35
##   Sampling weights variable                   sampling_ipw
## 
## Model Test User Model:
##                                               Standard      Scaled
##   Test Statistic                               182.920     107.676
##   Degrees of freedom                                26          26
##   P-value (Chi-square)                           0.000       0.000
##   Scaling correction factor                                  1.699
##     Yuan-Bentler correction (Mplus variant)                       
## 
## Model Test Baseline Model:
## 
##   Test statistic                              1424.722     838.746
##   Degrees of freedom                                55          55
##   P-value                                        0.000       0.000
##   Scaling correction factor                                  1.699
## 
## User Model versus Baseline Model:
## 
##   Comparative Fit Index (CFI)                    0.885       0.896
##   Tucker-Lewis Index (TLI)                       0.758       0.780
##                                                                   
##   Robust Comparative Fit Index (CFI)                         0.911
##   Robust Tucker-Lewis Index (TLI)                            0.811
## 
## Loglikelihood and Information Criteria:
## 
##   Loglikelihood user model (H0)             -32255.562  -32255.562
##   Scaling correction factor                                  1.630
##       for the MLR correction                                      
##   Loglikelihood unrestricted model (H1)     -32164.102  -32164.102
##   Scaling correction factor                                  1.654
##       for the MLR correction                                      
##                                                                   
##   Akaike (AIC)                               64613.125   64613.125
##   Bayesian (BIC)                             64910.375   64910.375
##   Sample-size adjusted Bayesian (SABIC)      64748.335   64748.335
## 
## Root Mean Square Error of Approximation:
## 
##   RMSEA                                          0.049       0.035
##   90 Percent confidence interval - lower         0.042       0.030
##   90 Percent confidence interval - upper         0.056       0.041
##   P-value H_0: RMSEA <= 0.050                    0.580       1.000
##   P-value H_0: RMSEA >= 0.080                    0.000       0.000
##                                                                   
##   Robust RMSEA                                               0.050
##   90 Percent confidence interval - lower                     0.039
##   90 Percent confidence interval - upper                     0.061
##   P-value H_0: Robust RMSEA <= 0.050                         0.469
##   P-value H_0: Robust RMSEA >= 0.080                         0.000
## 
## Standardized Root Mean Square Residual:
## 
##   SRMR                                           0.035       0.035
## 
## Parameter Estimates:
## 
##   Standard errors                             Sandwich
##   Information bread                           Observed
##   Observed information based on                Hessian
## 
## Latent Variables:
##                    Estimate  Std.Err  z-value  P(>|z|)   Std.lv  Std.all
##   RI_att =~                                                             
##     att_cbcl_w0       1.000                               0.537    0.607
##     att_cbcl_w1       1.000                               0.537    0.581
##     att_cbcl_w2       1.000                               0.537    0.571
##   RI_scr =~                                                             
##     screentm_rd_w0    1.000                               0.398    0.408
##     screentm_rd_w1    1.000                               0.398    0.407
##     screentm_rd_w2    1.000                               0.398    0.399
##   l_att_w0 =~                                                           
##     att_cbcl_w0       1.000                               0.704    0.795
##   l_att_w1 =~                                                           
##     att_cbcl_w1       1.000                               0.752    0.814
##   l_att_w2 =~                                                           
##     att_cbcl_w2       1.000                               0.773    0.821
##   l_scr_w0 =~                                                           
##     screentm_rd_w0    1.000                               0.892    0.913
##   l_scr_w1 =~                                                           
##     screentm_rd_w1    1.000                               0.893    0.913
##   l_scr_w2 =~                                                           
##     screentm_rd_w2    1.000                               0.914    0.917
## 
## Regressions:
##                    Estimate  Std.Err  z-value  P(>|z|)   Std.lv  Std.all
##   l_scr_w1 ~                                                            
##     l_scr_w0         -0.014    0.059   -0.235    0.814   -0.014   -0.014
##   l_scr_w2 ~                                                            
##     l_scr_w1         -0.083    0.070   -1.177    0.239   -0.081   -0.081
##   l_att_w1 ~                                                            
##     l_att_w0          0.148    0.073    2.024    0.043    0.138    0.138
##   l_att_w2 ~                                                            
##     l_att_w1          0.167    0.070    2.366    0.018    0.162    0.162
##   l_scr_w1 ~                                                            
##     l_att_w0          0.009    0.062    0.141    0.888    0.007    0.007
##   l_scr_w2 ~                                                            
##     l_att_w1          0.086    0.065    1.315    0.189    0.071    0.071
##   l_att_w1 ~                                                            
##     l_scr_w0          0.072    0.037    1.943    0.052    0.086    0.086
##   l_att_w2 ~                                                            
##     l_scr_w1          0.030    0.039    0.767    0.443    0.035    0.035
##   RI_att ~                                                              
##     Age_w0_z         -0.035    0.018   -1.981    0.048   -0.065   -0.065
##     Gender            0.094    0.018    5.173    0.000    0.174    0.175
##     abepstrat_w0     -0.035    0.018   -1.983    0.047   -0.066   -0.065
##     mnstlvl_4ct_w0   -0.001    0.019   -0.068    0.946   -0.002   -0.002
##     Mtrnl_DIAG_ANY    0.191    0.020    9.375    0.000    0.356    0.331
##   RI_scr ~                                                              
##     Age_w0_z          0.036    0.020    1.834    0.067    0.090    0.090
##     Gender            0.028    0.019    1.433    0.152    0.069    0.070
##     abepstrat_w0      0.038    0.019    2.006    0.045    0.096    0.095
##     mnstlvl_4ct_w0    0.053    0.020    2.639    0.008    0.132    0.132
##     Mtrnl_DIAG_ANY    0.067    0.019    3.539    0.000    0.169    0.158
## 
## Covariances:
##                         Estimate  Std.Err  z-value  P(>|z|)   Std.lv  Std.all
##  .RI_att ~~                                                                  
##    .RI_scr                -0.008    0.022   -0.336    0.737   -0.039   -0.039
##   l_att_w0 ~~                                                                
##     l_scr_w0               0.054    0.027    1.984    0.047    0.086    0.086
##  .l_att_w1 ~~                                                                
##    .l_scr_w1               0.085    0.032    2.692    0.007    0.129    0.129
##  .l_att_w2 ~~                                                                
##    .l_scr_w2               0.069    0.036    1.920    0.055    0.099    0.099
##   Age_w0_z ~~                                                                
##     Gender                -0.035    0.027   -1.275    0.202   -0.035   -0.034
##     abepstrat_w0           0.030    0.026    1.126    0.260    0.030    0.030
##     Mtrnl_DIAG_ANY         0.043    0.024    1.769    0.077    0.043    0.046
##     mnstlvl_4ct_w0        -0.072    0.026   -2.732    0.006   -0.072   -0.072
##   Gender ~~                                                                  
##     abepstrat_w0           0.043    0.025    1.713    0.087    0.043    0.043
##     Mtrnl_DIAG_ANY        -0.036    0.024   -1.498    0.134   -0.036   -0.038
##     mnstlvl_4ct_w0         0.090    0.026    3.414    0.001    0.090    0.090
##   abepstrat_w0 ~~                                                            
##     Mtrnl_DIAG_ANY        -0.037    0.023   -1.614    0.107   -0.037   -0.040
##     mnstlvl_4ct_w0         0.233    0.025    9.344    0.000    0.233    0.238
##   minstlevel_4cat_w0 ~~                                                      
##     Mtrnl_DIAG_ANY        -0.005    0.024   -0.213    0.831   -0.005   -0.005
## 
## Intercepts:
##                    Estimate  Std.Err  z-value  P(>|z|)   Std.lv  Std.all
##    .att_c_0 (iat1)   -0.145    0.022   -6.571    0.000   -0.145   -0.163
##    .att_c_1 (iat2)   -0.136    0.025   -5.401    0.000   -0.136   -0.147
##    .att_c_2 (iat3)   -0.071    0.030   -2.414    0.016   -0.071   -0.076
##    .scrn__0 (ist1)   -0.011    0.026   -0.403    0.687   -0.011   -0.011
##    .scrn__1 (ist2)   -0.024    0.030   -0.797    0.425   -0.024   -0.024
##    .scrn__2 (ist3)   -0.029    0.037   -0.770    0.441   -0.029   -0.029
## 
## Variances:
##                    Estimate  Std.Err  z-value  P(>|z|)   Std.lv  Std.all
##    .RI_att            0.247    0.035    6.974    0.000    0.856    0.856
##    .RI_scr            0.147    0.035    4.187    0.000    0.930    0.930
##     l_att_w0          0.495    0.040   12.281    0.000    1.000    1.000
##    .l_att_w1          0.550    0.043   12.837    0.000    0.972    0.972
##    .l_att_w2          0.581    0.044   13.343    0.000    0.971    0.971
##     l_scr_w0          0.795    0.043   18.294    0.000    1.000    1.000
##    .l_scr_w1          0.797    0.048   16.642    0.000    1.000    1.000
##    .l_scr_w2          0.827    0.052   16.017    0.000    0.990    0.990
##     Age_w0_z          1.005    0.028   36.327    0.000    1.005    1.000
##     Gender            1.010    0.005  199.247    0.000    1.010    1.000
##     abepstrat_w0      0.967    0.040   24.119    0.000    0.967    1.000
##     mnstlvl_4ct_w0    0.995    0.025   39.639    0.000    0.995    1.000
##     Mtrnl_DIAG_ANY    0.865    0.021   41.825    0.000    0.865    1.000
##    .att_cbcl_w0       0.000                               0.000    0.000
##    .att_cbcl_w1       0.000                               0.000    0.000
##    .att_cbcl_w2       0.000                               0.000    0.000
##    .screentm_rd_w0    0.000                               0.000    0.000
##    .screentm_rd_w1    0.000                               0.000    0.000
##    .screentm_rd_w2    0.000                               0.000    0.000
## 
## R-Square:
##                    Estimate
##     RI_att            0.144
##     RI_scr            0.070
##     l_att_w1          0.028
##     l_att_w2          0.029
##     l_scr_w1          0.000
##     l_scr_w2          0.010
##     att_cbcl_w0       1.000
##     att_cbcl_w1       1.000
##     att_cbcl_w2       1.000
##     screentm_rd_w0    1.000
##     screentm_rd_w1    1.000
##     screentm_rd_w2    1.000
```

```
parameterEstimates(sem_ri_clpm_screen_AttCBCL_adjust, ci = TRUE, level = 0.95, boot.ci.type = "perc", standardized = TRUE) %>% 
  filter(op == "~") %>% 
  dplyr::select('Regressions'=lhs, Indicator=rhs, B=est, SE=se, Z=z, 'p-value'=pvalue, Beta=std.all) %>% 
  kable(digits = 3, format="pandoc", caption="Regression coeficients from CLPM")
```

Regression coeficients from CLPM

| Regressions | Indicator | B | SE | Z | p-value | Beta |
| --- | --- | --- | --- | --- | --- | --- |
| l\_scr\_w1 | l\_scr\_w0 | -0.014 | 0.059 | -0.235 | 0.814 | -0.014 |
| l\_scr\_w2 | l\_scr\_w1 | -0.083 | 0.070 | -1.177 | 0.239 | -0.081 |
| l\_att\_w1 | l\_att\_w0 | 0.148 | 0.073 | 2.024 | 0.043 | 0.138 |
| l\_att\_w2 | l\_att\_w1 | 0.167 | 0.070 | 2.366 | 0.018 | 0.162 |
| l\_scr\_w1 | l\_att\_w0 | 0.009 | 0.062 | 0.141 | 0.888 | 0.007 |
| l\_scr\_w2 | l\_att\_w1 | 0.086 | 0.065 | 1.315 | 0.189 | 0.071 |
| l\_att\_w1 | l\_scr\_w0 | 0.072 | 0.037 | 1.943 | 0.052 | 0.086 |
| l\_att\_w2 | l\_scr\_w1 | 0.030 | 0.039 | 0.767 | 0.443 | 0.035 |
| RI\_att | Age\_w0\_z | -0.035 | 0.018 | -1.981 | 0.048 | -0.065 |
| RI\_att | Gender | 0.094 | 0.018 | 5.173 | 0.000 | 0.175 |
| RI\_att | abepstrat\_w0 | -0.035 | 0.018 | -1.983 | 0.047 | -0.065 |
| RI\_att | minstlevel\_4cat\_w0 | -0.001 | 0.019 | -0.068 | 0.946 | -0.002 |
| RI\_att | Maternal\_DIAG\_ANY | 0.191 | 0.020 | 9.375 | 0.000 | 0.331 |
| RI\_scr | Age\_w0\_z | 0.036 | 0.020 | 1.834 | 0.067 | 0.090 |
| RI\_scr | Gender | 0.028 | 0.019 | 1.433 | 0.152 | 0.070 |
| RI\_scr | abepstrat\_w0 | 0.038 | 0.019 | 2.006 | 0.045 | 0.095 |
| RI\_scr | minstlevel\_4cat\_w0 | 0.053 | 0.020 | 2.639 | 0.008 | 0.132 |
| RI\_scr | Maternal\_DIAG\_ANY | 0.067 | 0.019 | 3.539 | 0.000 | 0.158 |

```
#Standardized
Paths_sem_ri_clpm_screen_AttCBCL_adjust <- standardizedsolution(sem_ri_clpm_screen_AttCBCL_adjust, ci = TRUE, level = 0.95) %>% 
  filter(op != "=~") %>% 
  mutate(
    Beta_CI = sprintf("%.3f (%.3f to %.3f)", est.std, ci.lower, ci.upper),
    SE = sprintf("%.3f", se),
    Z = sprintf("%.3f", z),
    pvalue = sprintf("%.3f", pvalue),
    Beta = sprintf("%.3f", est.std)
  ) %>%
  dplyr::select('Regressions' = lhs, Indicator = rhs, Beta_CI, SE, Z, 'p-value' = pvalue, Beta)


Paths_sem_ri_clpm_screen_AttCBCL_adjust <- Paths_sem_ri_clpm_screen_AttCBCL_adjust[3:44,]
Paths_sem_ri_clpm_screen_AttCBCL_adjust <- Paths_sem_ri_clpm_screen_AttCBCL_adjust[-2:-7,]
Paths_sem_ri_clpm_screen_AttCBCL_adjust <- Paths_sem_ri_clpm_screen_AttCBCL_adjust[-10:-15,]
Paths_sem_ri_clpm_screen_AttCBCL_adjust <- Paths_sem_ri_clpm_screen_AttCBCL_adjust[-23:-30,]

wb <- loadWorkbook("ADHD_Screentime_Results.xlsx")
addWorksheet(wb, sheetName = "Adj RI-CLPM Screen-AttCBCL")
writeData(wb, sheet = "Adj RI-CLPM Screen-AttCBCL", x = Paths_sem_ri_clpm_screen_AttCBCL_adjust)
saveWorkbook(wb, file = "ADHD_Screentime_Results.xlsx", overwrite = TRUE)
```
